# A stochastic, individual-based model for the evaluation of the impact of non-pharmacological interventions on COVID-19 transmission in Slovakia

**DOI:** 10.1101/2020.05.11.20096362

**Authors:** Miroslav Gasparek, Michal Racko, Michal Dubovsky

## Abstract

The COVID-19 pandemic represents one of the most significant healthcare challenges that humanity faces. We developed a stochastic, individual-based model of transmission of COVID-19 in Slovakia. The proposed model is based on current clinical knowledge of the disease and takes into account the age structure of the population, distribution of the population into the households, interactions within the municipalities, and interaction among the individuals travelling between municipalities. Furthermore, the model incorporates the effect of age-dependent severity of COVID-19 and realistic trajectories of patients through the healthcare system. We assess the impact of the governmental non-pharmacological interventions, such as population-wide social distancing, social distancing within specific subsets of population, reduction of travel between the municipalities, and self-quarantining of the infected individuals. We also evaluate the impact of relaxing of strict restrictions, efficacy of the simple state feedback-based restrictions in controlling the outbreak, and the effect of superspreaders on the disease dynamics. Our simulations show that non-pharmacological interventions reduce the number of infected individuals and the number of fatalities, especially when the social distancing of particularly susceptible subgroups of the population is employed along with case isolation.

## Introduction

The outbreak of COVID-19 has become the most significant global health crisis of the 21^st^ century. As of 15th April, the Severe Acute Respiratory Syndrome Coronavirus 2 (SARS-CoV-2) has caused approximately two million confirmed cases of the disease and more than 125,000 deaths [1]. Currently, there is no known effective and definitive treatment available, although there are several promising antiviral drug candidates, such as chloroquine [2], Favipiravir [3], and Remdesivir [4]. Furthermore, there are multiple COVID-19 vaccines in the human clinical trials, such as mRNA-1273 by Moderna Therapeutics [5], Ad5-nCoV by CanSino Biologics [6], and ChAdOx1 nCoV-19 by University of Oxford and AstraZeneca [7] [8]. Nevertheless, it is expected that these vaccines will not be available earlier than in 12 months [9].

The world has not faced a disease of such lethality without access to the vaccine since the pandemics of H1N1 influenza (also known as “The Spanish flu”) [10]. Therefore, to contain the present COVID-19 pandemic, it is necessary to resort to Non-Pharmaceutical Interventions (NPI). These are measures leading to the reduction of the contacts between individuals, hence limiting the spread of the disease [10] [11]. In principle, there are two possible approaches to NPI [12]:

1. **Suppression**. In this strategy, the aim is to reduce the reproduction number of the disease, R (the average number of cases generated by an infectious individual in the population of fully susceptible people [13]) below 1.0. This would reduce the numbers of new cases to low levels like in the cases of SARS or Ebola [10] and eliminate the human-to-human transmission [10]. The main challenge associated with this approach is that the stringent NPI (such as social distancing and closure of schools) have to stay in place for a significant period of time until a vaccine is developed or while the virus is circulating in the human population [10]. Such NPI have a negative impact on economics and social cohesion.
2. **Mitigation**. In this strategy, the aim is not to completely avoid the transmission of the disease, but rather to utilize the NPI to reduce the load on the healthcare systems (“flatten the curve”) and to protect the members of the society vulnerable to the disease in a targeted fashion [10]. In this case, it is assumed that the immunity is built up throughout the epidemic, leading to a decrease in the number of cases and low transmission rates [10].

To contain the outbreak of COVID-19 while minimizing negative impacts on the health of the individuals, pressure on healthcare systems, and economic damage, it is necessary for governments to implement appropriate restrictive measures. To evaluate the impact of these measures, it is essential to utilize methods of mathematical and computational modelling that can inform the decisions of the key stakeholders. In this work, we develop a stochastic, individual-based model of COVID-19 transmission in Slovakia. We take into account demographic and geographic distribution of the population, daily migration within Slovakia, and periodic contacts among individuals, e.g., in their households. Our model is inspired by the state-of-art models published for the modelling of COVID-19 spread, such as [10]. The model can be used to evaluate the time evolution of the number of individuals infected by SARS-CoV-2, hospitalized individuals, individuals suffering from severe cases of COVID-19 that require critical care in Intensive Care Unit (ICU) or, equivalently, lung ventilation, and to estimate the number of potential fatalities. Furthermore, we evaluate the effect of different NPI on these counts and hence evaluate their effectiveness in containing the disease. We model the impact of various strategies on the containment of COVID-19 spread [10] [14] [15]. In our work, we use the data on the distribution of inhabitants in Slovak municipalities, data on migration between municipalities within Slovakia, the counts of individuals diagnosed with COVID-19 on 11th April 2020 provided by Slovak public health authorities, and the data from COVID-19 Mobility Trends Reports provided by Apple Inc. [16].

This work extends and complements the previous work on modelling COVID-19 outbreak in Slovakia that has been done by the Slovak Institute of Healthcare Policy (IZP), which utilized a deterministic Susceptible-Infected-Recovered (SIR) model with migration [17]. Our work provides insights into potential scenarios associated with COVID-19 outbreak in Slovakia, especially in terms of evaluation of the load on the number of ICU needed, which are critical for the successful management of the severe cases of COVID-19 [18] [19]. Our framework also offers the opportunity to model the more targeted and sophisticated NPI, such as inclusion of the effect of testing, utilization of intermittent or feedback-based NPI, and social distancing of the specific age sub-groups. Consequently, our model represents a *scenario-based projection* [20] that aims to offer a tool allowing public authorities to make more informed and effective selection of the appropriate NPI. Our contributions are:

- Development of a stochastic, individual-based model of COVID-19 spread in Slovakia.
- Evaluation of the effect of non-pharmaceutical interventions (NPI) and evaluation of their impact on the demands on the healthcare system capacities.

## Prior work

The most consequential models of COVID-19 spread were published by Ferguson et al. [10] [21]. These models describe the spread of COVID-19 across the UK and US [10] and globally [21]. The models are based on the original model of the spread of pandemic influenza by Halloran, et al. [11]. In these modelling frameworks, individuals reside together in households, attend schools, workplaces, and move in the wider community, where contacts between the individuals occur [10]. The model described in [10] utilizes detailed Census data to approximate these phenomena and to model the interactions between the individuals. While reproducing this model is not feasible due to the absence of the required data, its conceptual importance and some key aspects of this work (e.g., hospitalization, ICU requirements and various NPI) informed our work. The model assumes that the transmissions of the disease occur between the susceptible and infectious individuals at all the aforementioned places (home, school, workplace, or randomly in the community; where the last type of transmission depends on the spatial distance). The importance of this model lies in the fact that it allows for the evaluation of the impact of *mitigation* and *suppression* [10] on the spread of the disease. The brief summary of these strategies can be found in [10] and [22]. Ferguson et al. evaluated the impact of different NPI utilized in mitigation or suppression strategy, either individually or in combination. While this model has been criticised [23], we believe that the critique is not based on substantial grounds, as the authors do not provide research results refuting the work in [10].

The work presented in [10] has been expanded; the model of COVID-19 spread was applied at the global scale and evaluated the impact of various NPI and their combinations in 202 countries [24]. Another extension of work in [10] has been presented in [22], where Flaxman et al. evaluated the impact of NPI on COVID-19 in 11 European countries, suggesting that these measures were effective in reducing the detrimental health impact of the disease [22]. Other COVID-19-related modelling includes the work by Kucharski et al., in which the transmission of COVID-19 disease in Wuhan was modelled using SEIR stochastic transmission dynamical model fitted to multiple public data sets [14] and work of Hellewell et al. that focused on the evaluation of the impact of isolation of cases and contacts on the spread of COVID-19 epidemics [25]. There have been numerous publications focused on modelling of different aspects of COVID-19 dynamics: estimation of the epidemiological parameters [15] [26] [27], deterministic and stochastic modelling of the COVID-19 outbreak and its control [28] [29] [30], spatial modelling [31], modelling of the disease spread on heterogeneous complex network [32], and modelling of the optimal lock-down scenarios from the point of view of the optimal control theory [33].

## Methods

### Model overview

We developed a stochastic, individual-based transmission model of COVID-19 outbreak based mainly on the work in [10] [29] [34]. The model captures the features of the COVID-19, which are important in this disease, as outlined in [29]:

- A significant proportion of asymptomatic carriers [35]
- Unprecedented public campaigns encouraging self-isolation after the onset of symptoms [36]

We devised the stochastic model with the following states, assuming that the total population is fixed (Figure 1):

- *S*, **Susceptible**: Individuals that have not contracted the disease and are not immune.
- *E*, **Exposed**: Individuals that have been exposed to the disease, have been infected, but are not yet infectious. These individuals are in the latent period.
- *I_A_*, **Infected asymptomatic**: Individuals that have contracted the disease and are infectious period, but their disease has a mild course and therefore are not traced or self-isolated.
- *I_S_*, **Infected symptomatic**: Individuals that have contracted the disease, are infectious and display the disease symptoms.
- *H*, **Hospitalized**: Individuals that have contracted the disease, are symptomatic and their course of the disease requires hospitalization.
- *H_C_*, **Hospitalized in ICU**: Individuals that have contracted the disease, are symptomatic and their course of the disease requires ICU.
- *R*, **Recovered**: Individuals that had contracted the disease recovered.
- *D*, **Dead**: Individuals that had contracted the disease, were symptomatic, their course of the disease required ICU, and did not survive.

**Figure 1.**
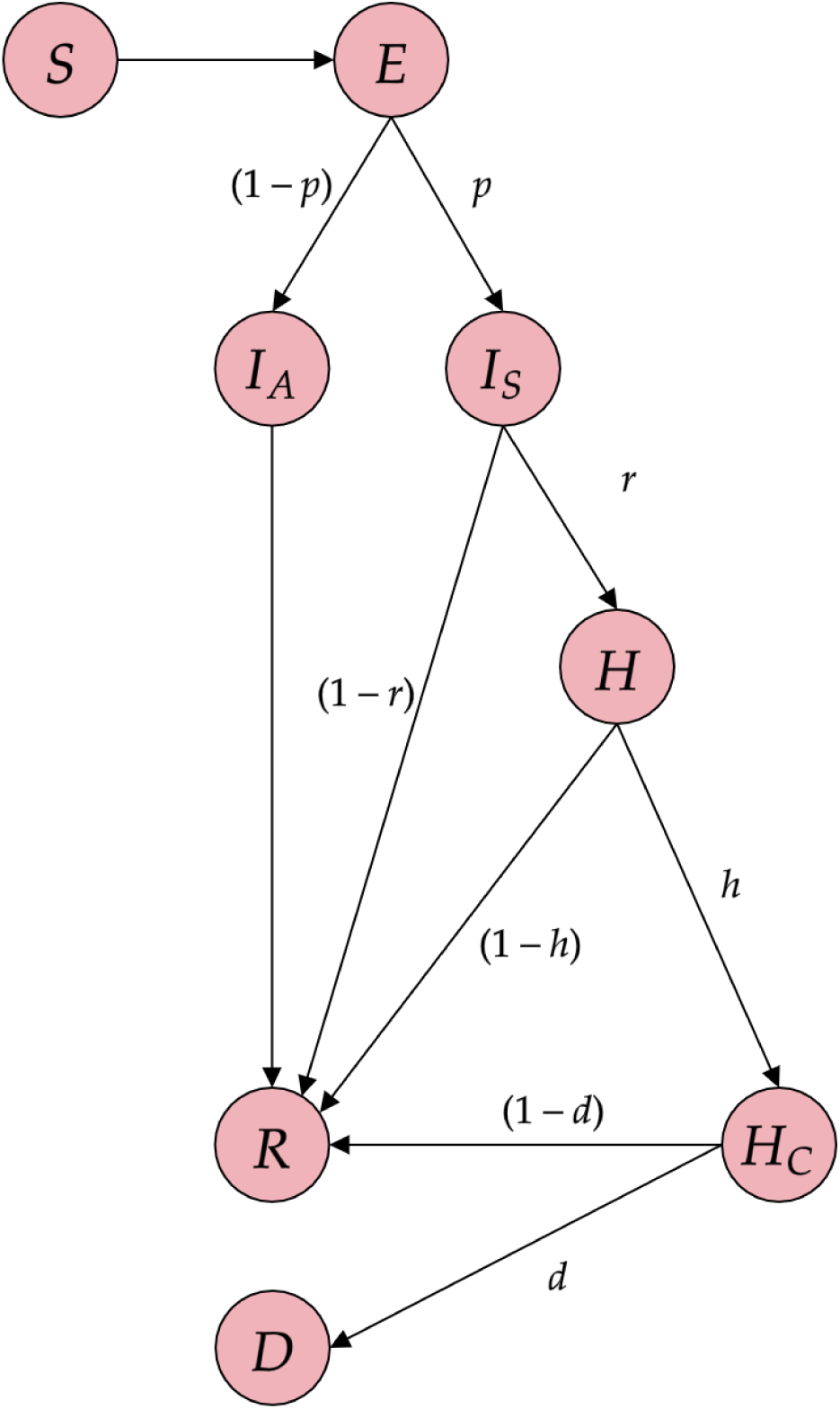
The schematics and overview of the states in the stochastic model of COVID-19 epidemics. Red circles represent the states: Susceptible *(S*), Exposed (*E*), Infected Asymptomatic *(I_A_)*, Infected Symptomatic *(I_S_*), Hospitalized (*H*), Hospitalized requiring ICU (*H_C_*), Recovered (*R*) and Dead (*D*). Letters next to the arrows denote the transition probabilities to the given states.

### Distribution into households and interactions between the individuals

In the model, individuals interact within the households, in the municipalities where they live, and in the other municipalities to which they travel. Within households, individuals interact with the same individuals at each time step (every day). The numbers and age of the individuals in each household are drawn from the discrete probability distribution with probability mass function fitted to the data from Statistics on Income and Living Conditions provided by Eurostat [37] (Figure S5). The comparison of the real household data with the simulated data is in the Supplementary Information (Figure S1). Within their own municipality, individuals interact in a random fashion with co-inhabitants of this municipality, and with the individuals travelling to their home municipality.

Travelling between the municipalities is described by the data from the Origin-Destination Matrix (ODM) [38], *M*, where *M_i,j_* is the average number of people per day that travel from the municipality *j* to municipality *i*. To add the daily variation in the travelling between the municipalities, we assume that on every time step of the simulation (1 day), a portion of the population with size *S_ij_ ~ Poisson(λ* = *M_i,j_*) from each municipality *j* is selected and interacts randomly with the population in the municipality *i*. The proportion of infected cases travelling to the city *i* from *j* is equal to the proportion of infected cases in the municipality *j* on the given day, excluding those who are hospitalized or quarantined. Furthermore, we assume that the interaction within the municipalities and between municipalities captures all the travel, school attendance, work-related movement, and other types of social interaction (based on the recommendation of IZP).

### Initialization

At the start of the simulation, the population in all locations is initialized as susceptible (S), with the exception of a small subset of the population that is initialized as exposed (E). No individuals are initially present in the other states, as the numbers of Hospitalized, Hospitalized in ICU, Recovered and Dead individuals as of 11th April 2020, which we take as the starting day of the simulation, were negligible with respect to the total population of Slovakia. Furthermore, their impact on the disease dynamics was negligible, as we assumed that these individuals were isolated and cannot spread the disease further. We assumed that the initial locations of the exposed individuals that have been infected were as published on 11th April 2020. The number of infected (exposed) individuals at the start of the simulation was set to be 729 in all simulations, which corresponds to the data available from the IZP. We note that because of the relatively low level of testing in Slovakia and high number of asymptomatic infections, this initial number of exposed individuals almost certainly underestimates the true number of infected individuals. However, the slow spread of the disease across Slovakia that has been observed so far justifies these numbers.

### Age-dependent stratification of the severity of COVID-19 cases

We also took into account the age-dependent severity of the disease, utilizing the data from Ferguson et al. [10] and Verity et al. [39]. The age-dependent transition probabilities between the different states are shown in Table 1. Such stratification of the COVID-19 severity is very important, as it has been clinically observed that the proportion of the severe cases of the disease is greater for the elderly. Infection Fatality Ratio in Table 1 is provided as the proportion of infected individuals who die as a consequence of the disease. We assumed that only the individuals that require ICU can die (transition to “Dead” state). It is known that even access to the ICU does not ensure that the individuals with severe cases of the disease survive [40]. However, in the case of the overwhelmed healthcare system, also the individuals that otherwise would not require ICU care could be in the risk of dying. Consequently, our estimate of the number of fatalities is rather optimistic.

**Table 1.**
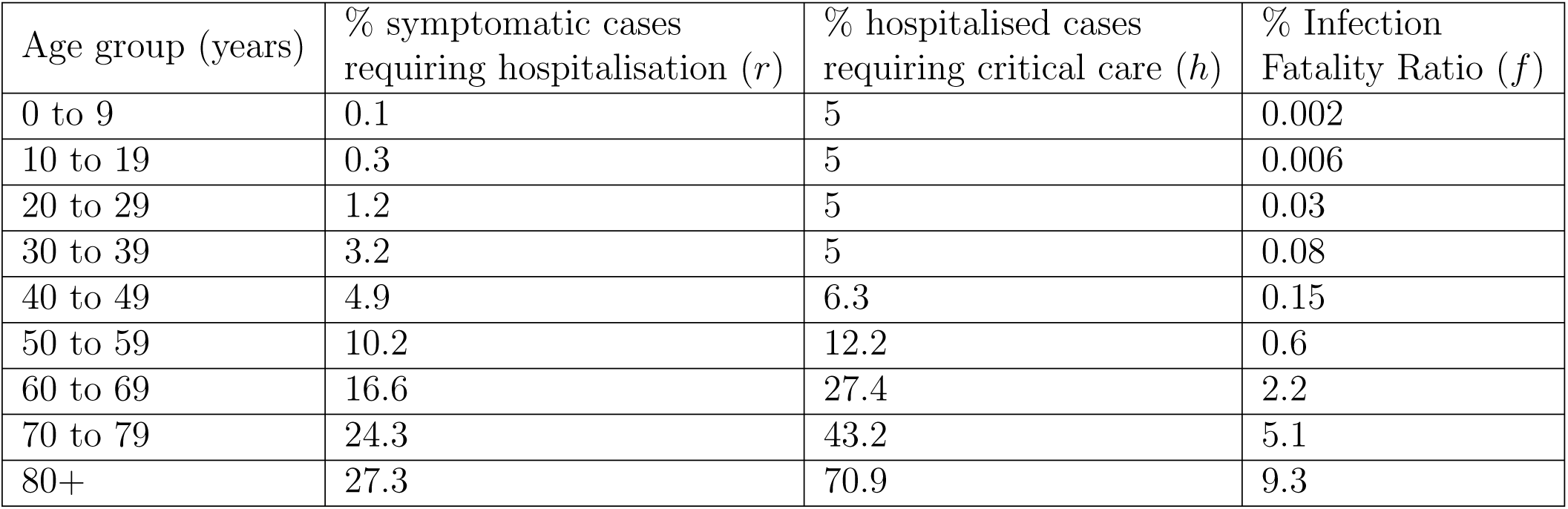
Estimates of the age-dependent severity of the disease adopted from Verity et al. [39]. Columns display the values of transition probabilities between the states for each of the age groups.

### Transmission dynamics

Transmission events occur through interaction between the susceptible and infectious individuals, either in the households, through the random contacts in their home municipalities, or through the random interactions during the travel to the other municipalities [10]. The incubation period (i.e., time between infection and onset of symptoms) was drawn from the Normal distribution with a mean of 5.1 days and Variance 1 *Day*^2^. We assumed that an individual does not become infectious until the onset of the symptoms, based on the fact that the infectiousness of the individual before the onset of the symptoms is significantly lower [41]. We also assumed that 40% of the individuals are asymptomatic or have a very mild course of the disease, which means that they do not display symptoms of COVID-19, but they can still spread the disease based on the assumptions in [10] during the period of infectiousness. Hence, we assume that only 60% of patients that are infected will display the symptoms of COVID-19. Furthermore, we assumed that the duration of the infectiousness (i.e. the period during which the patient is infectious) is sampled from Normal distribution with a mean of 6.5 days and Variance of 1 *Day*^2^. While this value varies based on different sources, we believe that due to the increased public awareness and self-isolation measures, individuals either self-isolate themselves within that period after the onset of symptoms or experience severe course of the disease and are treated in the hospitals under the strict quarantine measures, where the possibility of spread is limited due to the extended use of the personal protection equipment [25].

The important parameter that governs the spread of epidemics is the basic reproduction number, *R*_0_. Based on the early growth rate in Wuhan [15] [34], we set *R*_0_ = 2.4 in the absence of any NPI. We assumed that the individuals who have been infected are immune to reinfection in the short term [10] [42], although more research is needed to confirm this assumption [43]. Time from the infection to hospitalization was sampled from Normal distribution with a mean of 10 days and Variance of 1.5 *Day*^2^ [10]. We assumed that if an individual is hospitalized and has a mild course of the disease, she spends the time in hospital that is drawn from Normal distribution with a mean of 8 days and Variance of 1 *Day*^2^ [10]. If an individual suffers from the severe course of the disease, she first spends time drawn from Normal distribution with a mean of 6 days and Variance of 1.5 *Day*^2^ in the normal hospital bed [10] and subsequently spends time drawn from Normal distribution with a mean of 10 days and Variance of 1 *Day*^2^ in the ICU [10] (Figure 2, Figure S9, Table 2.)

**Figure 2.**
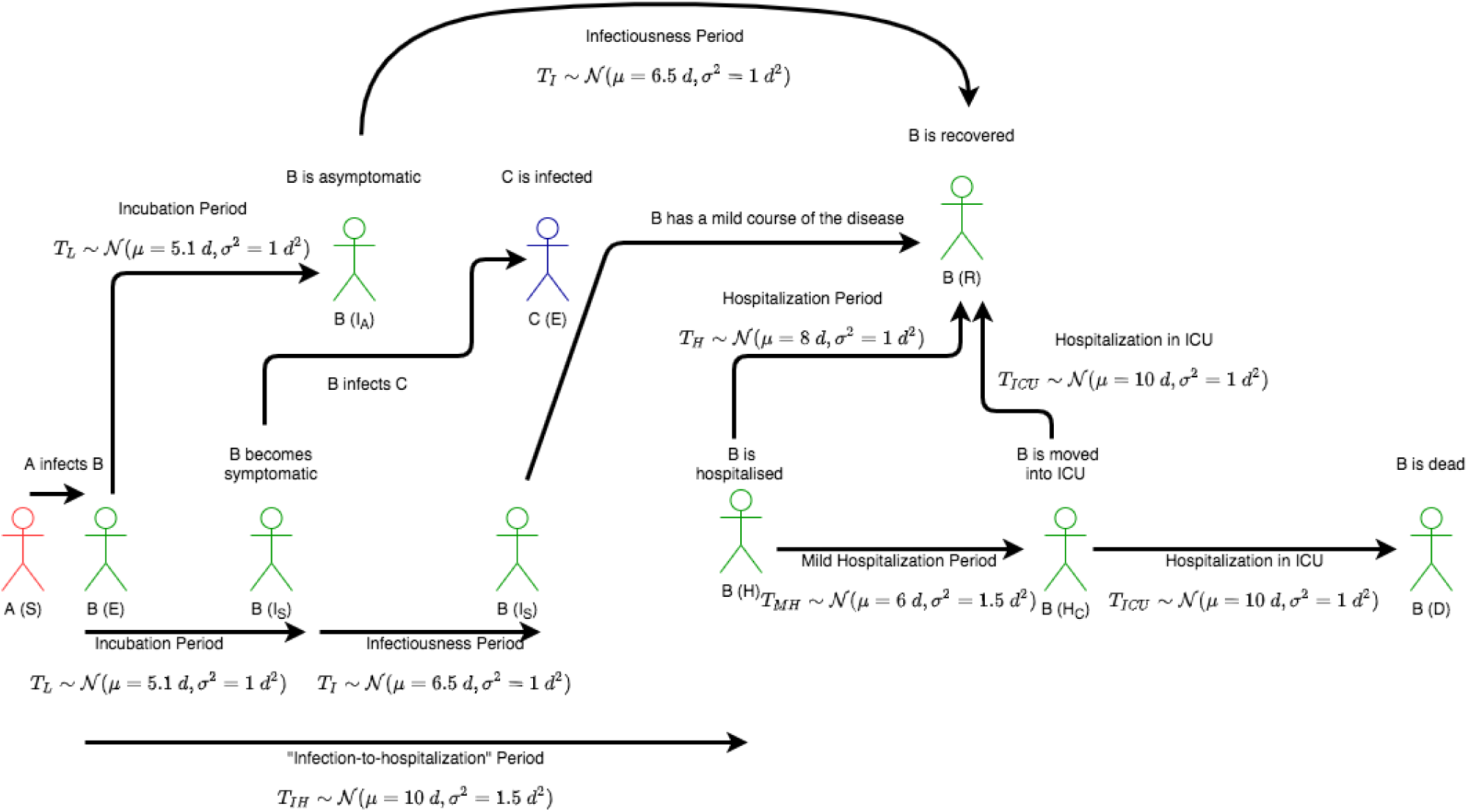
Schematics of the disease transmission process. Infected individual A infects susceptible individual B. After the incubation period, individual B becomes infectious and develops symptoms or has an asymptomatic course of the disease. During the period of infectiousness, infected individual B may infect susceptible individual C. After the infectiousness period, individual B either moves into the Recovered state or into the Hospitalized state. If individual B is hospitalized, he can either move into Hospitalized in ICU state or Recovered state. Finally, if the patient B is in ICU, the patient can move into Dead state or Recovered state.

**Table 2.** Parameters values and probability distributions used in the simulation of COVID-19 epidemics.

| Parameter | Distribution | Value |
| --- | --- | --- |
| Incubation period | $\mathcal{N}(\mu = 5.1 \text{ d}, \sigma^2 = 1 \text{ d}^2)$ | |
| Infectiousness duration | $\mathcal{N}(\mu = 6.5 \text{ d}, \sigma^2 = 1 \text{ d}^2)$ | |
| Infection-to-hospitalization period | $\mathcal{N}(\mu = 10 \text{ d}, \sigma^2 = 1.5 \text{ d}^2)$ | |
| Hospitalization period<br>in the mild case of the disease | $\mathcal{N}(\mu = 8 \text{ d}, \sigma^2 = 1 \text{ d}^2)$ | |
| Mild hospitalization period<br>in the case of a severe<br>course of the disease | $\mathcal{N}(\mu = 6 \text{ d}, \sigma^2 = 1.5 \text{ d}^2)$ | |
| ICU hospitalization period<br>in the case of a severe<br>course of the disease | $\mathcal{N}(\mu = 10 \text{ d}, \sigma^2 = 1 \text{ d}^2)$ | |
| Number of random contacts<br>outside the household<br>per day in unmitigated scenario | $Poisson(\lambda = 15)$ | |
| The probability that a<br>susceptible is infected<br>given that single infected<br>individual is in the household | | $\beta_{H0} = 0.064$ |
| Probability of infected<br>individual being<br>symptomatic | | $p = 0.60$ |
| Probability of infected<br>individual requiring<br>hospitalization | | Value of $r$ in Table 1 |
| Probability of hospitalized<br>individual requiring<br>hospitalization in ICU | | Value of $h$ in Table 1 |
| Probability of individual in ICU<br>dying from COVID-19 | | Value of $d = f/h$ in Table 1 |

### Modelling transmission events

Assuming that the members of each household meet daily, we assume that the probability of the transmission in the household on the given day is *β_H_* = *β_H_*_0_*n_I_*, where *n_I_* is the number of infected individuals in the household and *β_H_*_0_ = 0.064. This estimate is based on the estimates of the secondary attack rates [44] published in [45]. To model the transmission events outside the household, we evaluated the probability of transmission of the disease during the random contact between the infectious and susceptible individual, *β*, as follows: we defined the reproduction number *R* = [(1 + *r_T_)nβ* + *β_h_*] *τ*, where *τ* is the duration of the infectiousness period in days, *n* is the number of the random contacts that typical individual has per day, and *r_T_* is the ratio of all individuals travelling between the municipalities to the whole population. For each infected individual, we sampled *τ* from normal distribution, 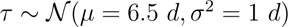, while *β* was drawn from the uniform distribution, 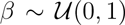 and *n* was drawn from the Poisson distribution, *n* ~ *Poisson(λ* = 15), where λ = 15, is the characteristic number of daily random contacts that an individual has in the absence of any restrictions. This value was partially justified by the average number of meetings recorded in Wuhan before the social distancing measures were imposed [46]. We computed the mean values of *τ* and *n*, re-computed the mean probability of transmission between infected and susceptible individuals as

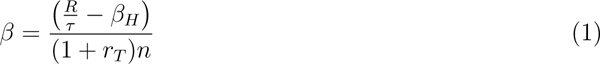

and we run the simulation again with the computed values, utilizing mean-field approximation [47].

### Simulations

We ran the simulations with the time step of 1 day for the various lengths of time vectors, until the steady-state distribution was reached or satisfactory insights into the behaviour of the system were obtained. The model was implemented in Python programming language and was optimized for running on GPU by using the CuPy package.

## Results

### A. Evaluating the spread of COVID-19 in Slovakia in the absence of NPI

First, we simulated the spread of COVID-19 in the absence of the social-distancing measures, assuming that the characteristic number of contacts among the individuals outside the household is *n* ~ *Poisson(λ* = 15). We initialized the simulation based on the published number and locations of infected individuals as described in the Methods section. Such a scenario corresponds to the unmitigated strategy, in which the disease is allowed to spread through the population so that the collective immunity can be built up. The simulation was conducted as described in the Methods section (Figure 3). The simulation suggests that the total number of the infected individuals is approx. 63% of the total population. The overall number of deaths nears 36,000, or around 0.65%, which is significantly lower than the adjusted Case Fatality Ratio (CFR) of 1.38% reported in [48] for China. The peak in the number of hospitalized occurs 56 days after the start of the simulation. The peak number of ICU beds required is approx. 21,000, which far exceeds the capacities of Slovak healthcare system (black dashed line in the Figure 3). These results suggest that the absence of the governmental action enforcing the social distancing would lead to a significant burden on the healthcare system and to the significant number of fatalities, especially considering that the current Slovak ICU capacity is only at around 500 ICU beds with ventilators [49], with the planned increase to 1000 ICU beds with ventilators.

**Figure 3.**
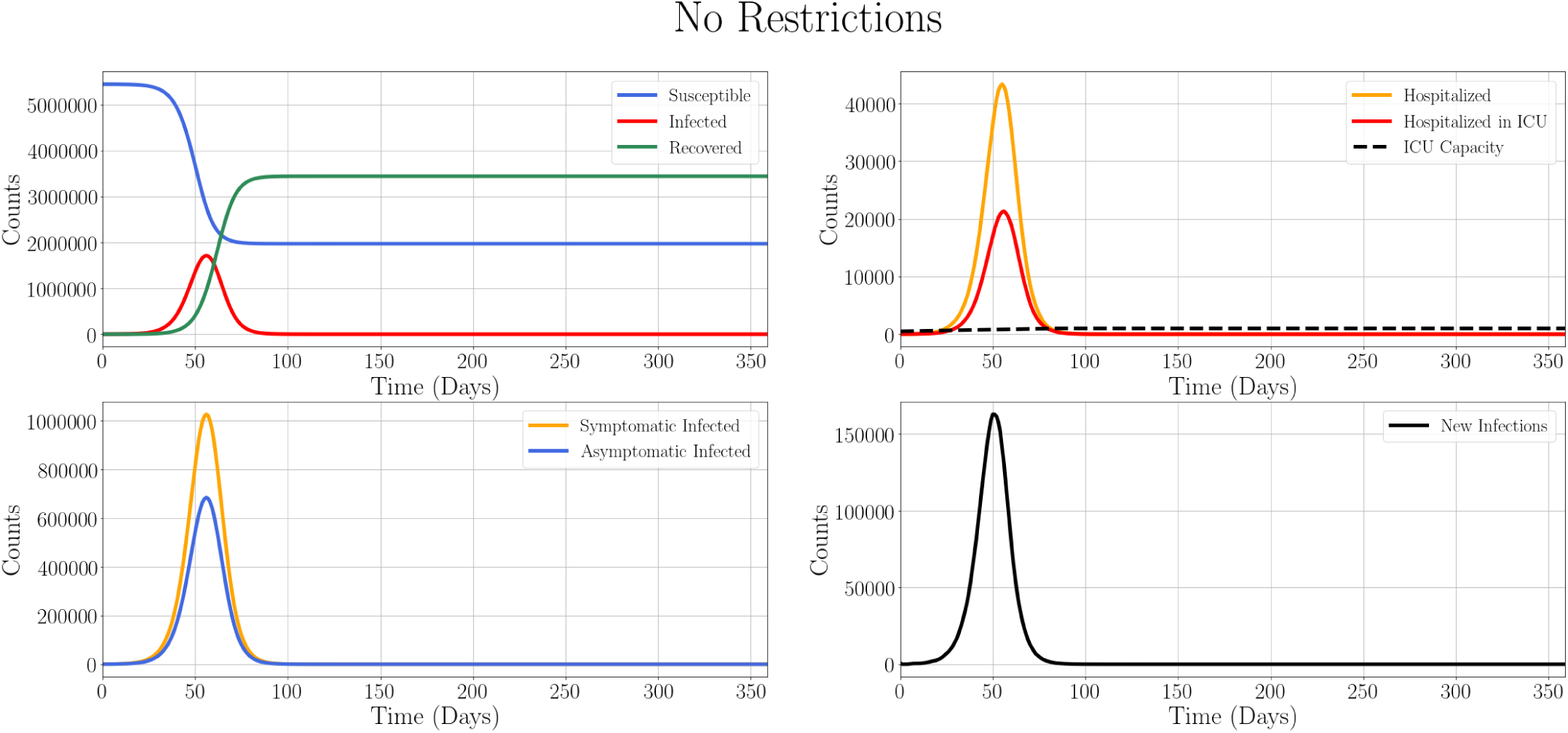
Time evolution of COVID-19 spread through Slovakia in the absence of social distancing and mobility restriction measures. (Upper Left) Counts of Susceptible (blue), Infected (red), and Recovered (green) individuals. (Upper Right) Numbers of individuals requiring hospitalization (orange) and hospitalization in ICU care (red) and the maximum ICU capacity (dashed black). (Lower Left) The time evolution of the counts of Infected Symptomatic individuals (yellow) and Infected Asymptomatic individuals (blue). (Lower Right) The time evolution of the daily new infections (black). The number of the hospitalized individuals peaks on Day 56 of the simulation.

To verify the results of our model, we compared the deterministic SEIR model [13] [50] (Figure S9) with the results obtained using this model without any NPI applied, with no household transmissions and no travel between the municipalities. We found a good agreement between both models.

### B. Estimation of the COVID-19 spread in the presence of the current NPI

After the first cases of the COVID-19 in Slovakia, strict social distancing and mobility restriction measures such as mandatory face mask wearing in the public spaces, closure of the schools, universities, churches, and non-essential businesses were imposed. To assess the impact of the decrease in mobility currently imposed on Slovakia, we utilized the Apple Mobility Trends Reports [16], which describes the relative mobility changes in terms of Driving, Walking, and Transit when compared to the baseline value from 13th January 2020 across the whole country. These data on the mobility reduction are shown in Table 3. While such data have significant limitations such as only taking into account the movement of users of the Apple devices and the baseline determined using one data point only, it is currently the best available mobility indicator that we have at our disposal.

**Table 3.** The relative change in driving, walking, and transit data. Adopted from Apple Mobility Trends Reports on 18th April 2020 [51].

| Sector | % Relative mobility change |
| --- | --- |
| Driving | -36% |
| Transit | -52% |
| Walking | -26% |

We utilized these data to infer the approximate reduction in the number of the random contacts. We transferred the reduction in “Walking” into the reduction in the characteristic number of random contacts that individuals typically have within their own municipalities. In other words, we assumed that for each individual, the number of random contacts within the municipality is *n* ~ *Poisson(λ* = 0.74 × 15). Similarly, based on the reduction in “Driving“, we estimated that the mobility of individuals between the municipalities was reduced to 64% of its typical value. This means that each entry of the Origin-Destination Matrix was multiplied by the factor 0.64.

One of the most significant social distancing measures that have been implemented was the *quarantining of the infected individuals*. To model effect of quarantine, we assumed that one day after the development of symptoms (i.e., after the end of incubation period 1 day) the symptomatic infectious individuals have the probability of disease transmission reduced to 20% of its original value for 14 days. Non-zero transmission probability accounts for the disobedience of the social distancing measures, or unintentional spread of the disease. We assumed that the probability of the transmission within the household did not change. We also stress that we did not consider the effect of the NPI such as wearing of the face masks and gloves in public, and physical social distancing in the public spaces, which can further significantly reduce the transmission rate of the disease.

We modelled the effect of the potential prolonged stringent NPI on the spread of the disease in Slovakia and the load on the healthcare capacities. We initialized the simulation as described in Methods section, assuming that NPI and reduction in mobility was in place for 240 days (roughly until the end of 2020). After this period, we assumed that the restrictions were completely lifted (Figure 4).

**Figure 4.**
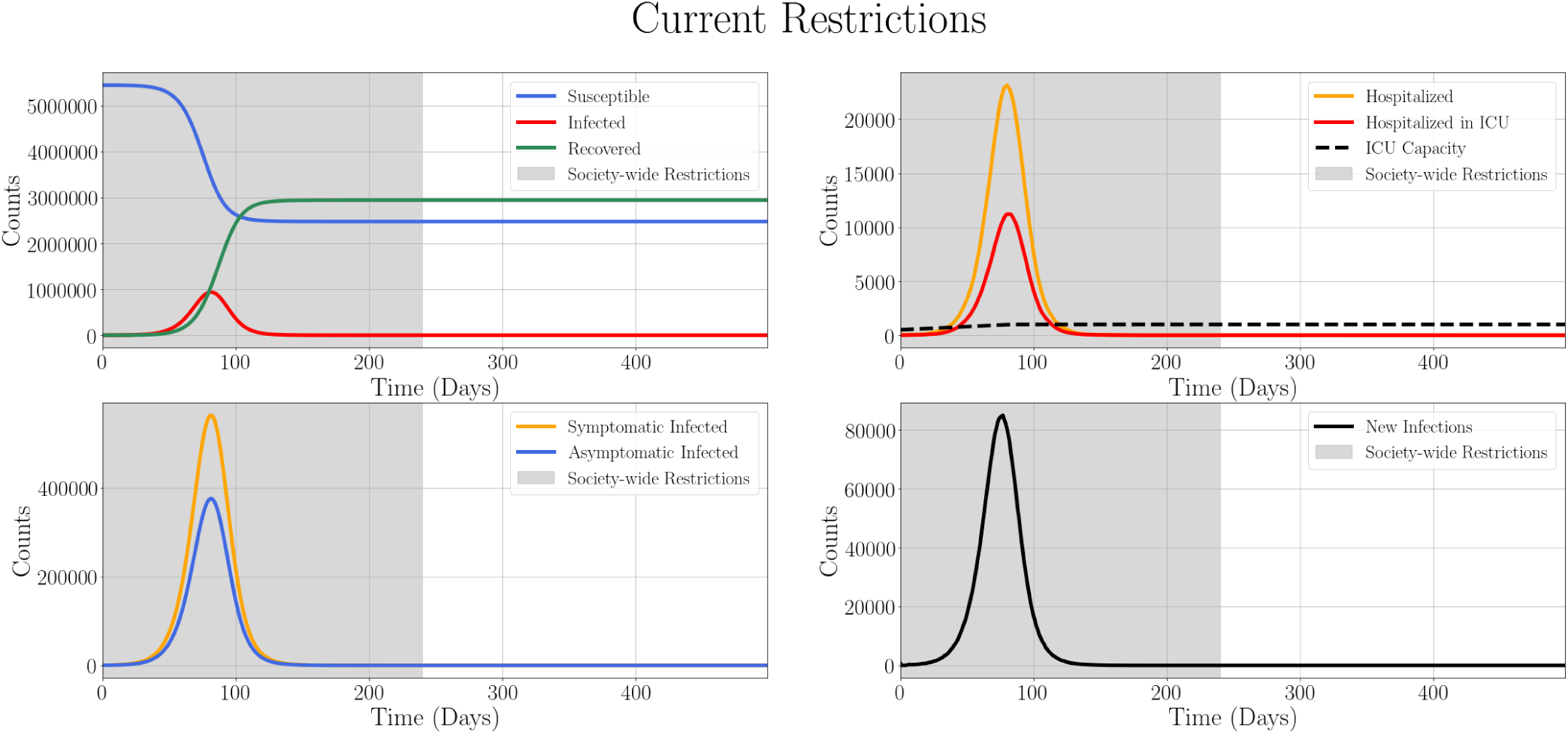
Time evolution of COVID-19 spread through Slovakia in the presence of the current estimated level of social distancing measures and mobility restrictions. The restrictions are introduced at the start of the simulation and are lifted 240 days later.(Upper Left) Counts of Susceptible (blue), Infected (red), and Recovered (green) individuals. (Upper Right) Numbers of individuals requiring hospitalization (orange), hospitalization in ICU (red) and the maximum ICU capacity available (dashed black). (Lower Left) The time evolution of the counts of Infected Symptomatic individuals (yellow) and Infected Asymptomatic individuals (blue). (Lower Right) The time evolution of the daily new infections (black). Grey shading of the plots depicts the period during which the NPI are in place. The number of the hospitalized individuals requiring ICU peaks on Day 80 of the simulation.

The simulation suggests that the total number of the infected individuals is approx. 54% of the total population. The peak in the number of hospitalized occurs 80 days after the start of the simulation. Although peak number of ICU beds needed is still above the expected number of ICU beds available at the end of June 2020, these requirements are reduced to 11,200, which is approximately a half of the number required in the case without any NPI applied. Furthermore, the peak in the number of ICU needed is significantly delayed. These results suggest the effectiveness of social distancing measures in decreasing the negative impact of COVID-19 on healthcare systems.

### C. Short-term strict NPI followed by the isolation of the vulnerable sub-populations provides better trade-off between the load on healthcare system and economic damage

While the strict NPI can reduce the burden on the healthcare systems and slow down the spread of the epidemics, implementation of NPI is associated with the negative impacts on the economy [12] and the social fabric [52]. Therefore, it is desirable to restart the post-epidemic economic activity as soon as possible. This restart can be facilitated by the implementation of the more targeted and fine-tuned social distancing measures for the vulnerable sub-groups, such as elderly, patients with the chronic respiratory or cardiovascular diseases, who often suffer from the more severe course of the disease [39]. To demonstrate the effect of such targeted measures, we simulated the implementation of the relatively strict NPI as described in Results, Section B, for 60 days. Subsequently, we released these strict NPI and we restricted the mobility of the elderly for the rest of the simulation (210 days). That is, the mobility of the population aged above 60 was restricted to 20% (Figure 5).

**Figure 5.**
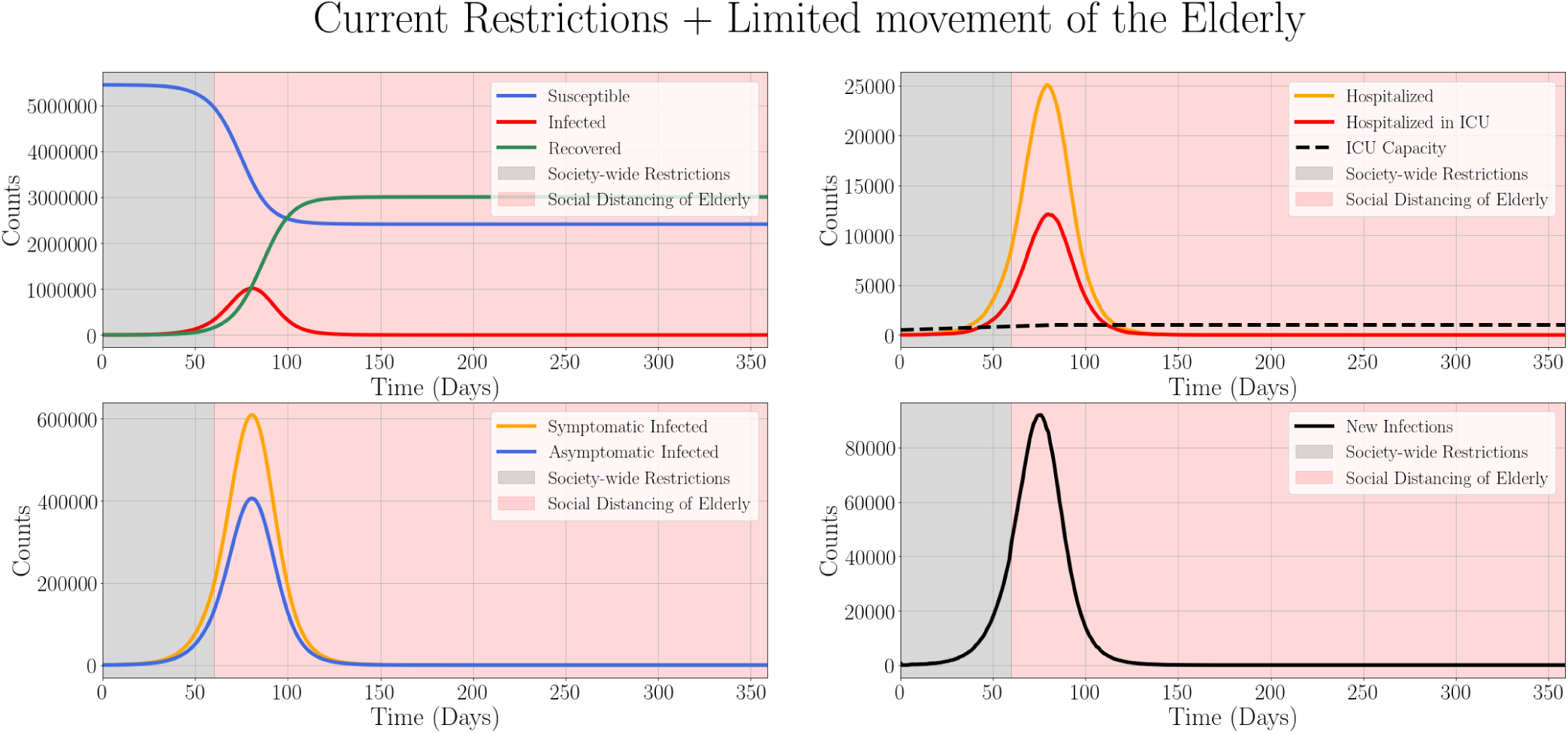
Time evolution of COVID-19 spread through Slovakia in the presence of the current estimated level of social distancing and mobility restrictions, and the subsequent social distancing of the elderly only. The mobility and social distancing restrictions are introduced at the start of the simulation and are lifted 60 days later. Subsequently, the mobility of the individuals aged above 60 is restricted until the end of the simulation. (Upper Left) Counts of Susceptible (blue), Infected (red), and Recovered (green) individuals. (Upper Right) Numbers of individuals requiring hospitalization (orange), hospitalization in ICU (red) and the maximum ICU capacity available (dashed black). (Lower Left) The time evolution of the counts of Infected Symptomatic individuals (yellow) and Infected Asymptomatic individuals (blue). (Lower Right) The time evolution of the daily new infections (black). Shadings of the plots depict the period during which the strict NPI are in place (grey) and social distancing of the elderly is in place (red). The number of the hospitalized individuals peaks on Day 79 of the simulation.

The results of the simulation show that the total number of the infected individuals is approx. 55% of the total population of the country. The peak in the number of hospitalized occurs 79 days after the start of the simulation. The peak number of ICU beds required is approx. 12,100. While this number is still considerably above the expected number of ICU beds available, the ICU requirements are significantly reduced when compared to the case of the unmitigated scenario. Furthermore, the strict social distancing measures that negatively impact the economy are in place only for 60 days, a considerably shorter period than the prolonged time for which the NPI were implemented in the previous scenario (Figure 4). This result shows that targeted social distancing of the vulnerable, but less economically active subgroups can play a significant role in the fast economic restoration, although it is important to consider the adverse effects of the social distancing on the psychological well-being and reduction of the compliance in the case of the long-term restrictions [52].

### D. Introduction of the strict restrictions followed by their full release does not prevent the subsequent spread of COVID-19

Recently, a so-called “lock-down scenario” has been reportedly discussed by some members of the Slovak government [53]. In this hypothetical scenario, all the citizens would be mandated to stay in their households, except of those who are essential for maintaining the fundamental functioning of the state apparatus, such as those employed in medical fields, armed forces, police, and logistics. We explored the potential impacts of this scenario on the spread of COVID-19, while neglecting the immense economic and social costs associated with this strategy. We initialized the simulation on 11th April 2020 as described in Methods section, with the number of contacts for the individuals travelling between the municipalities reduced to 74% of the non-restricted value. After 40 days, we introduced the “lock-down”: we abruptly reduced the number of random contacts outside the household from its typical characteristic value (15 random contacts/day) to 5% of this value. This accounts for both disobedience of the restrictions by the citizens, as well as the need for maintaining the basic logistical functions within the country. We assumed that the contacts in the households are unaffected during the “lock-down”. After 60 days, the strict restrictions were lifted, i.e., mobility was restored to 74% of the non-restricted value (Figure 6).

**Figure 6.**
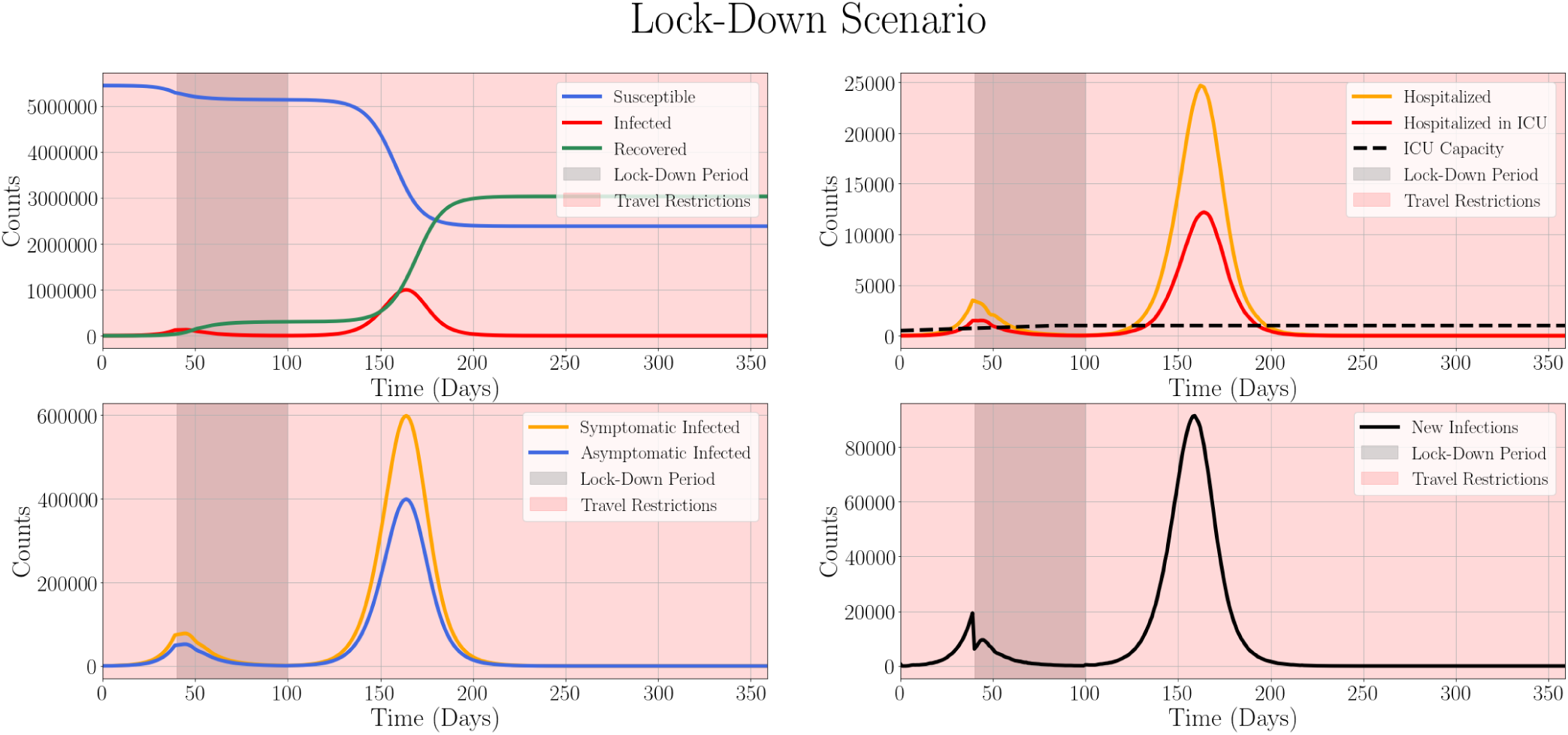
Time evolution of COVID-19 spread through Slovakia in the case of so-called “lock-down” scenario. Initially, the number of random contacts for the individuals travelling between the municipalities is 74% of the non-restricted value. The number of all random contacts between individuals is reduced on Day 40 of the simulation to 5% of its initial value. Mobility is reduced for 60 days and subsequently rises back to 100% afterwards until the end of the simulation, except for the number of contacts due to the travel between the municipalities, which rises to 74% of the unrestricted value. (Upper Left) Counts of Susceptible (blue), Infected (red), and Recovered (green) individuals. (Upper Right) Numbers of individuals requiring hospitalization (orange) and hospitalization in ICU care (red) and the maximum capacity of the ICU available (dashed black). (Lower Left) The time evolution of the counts of Infected Symptomatic individuals (yellow) and Infected Asymptomatic individuals (blue). (Lower Right) The time evolution of the daily new infections (black). Grey shading of the plots depicts the period during which the strict NPI are in place. The peak in the number of the hospitalized individuals requiring critical care occurs on Day 162.

It is clear that the introduction of the severe restrictions drive the number of new cases and requirements for hospitalizations and ICU care almost to zero. However, after such restrictions on mobility and social distancing measures are lifted, there is a rebound in the number of new cases of COVID-19 and the demands for ICU raises to more than 12,000, albeit this raise is delayed. This suggests that 60 days of almost total restriction of the movement in the public spaces would not eradicate the disease from the population, which might be due to the spread of the virus in the households [45] even without consideration of the possible import from abroad after the opening of the borders. We also investigated the effect of shorter (30 days) and extended (120 days) “lockdown” on the disease dynamics. The results (Figure S2) show that even 120 days of “lock-downs” do not lead to the complete eradication of the disease and the rebound occurs after the restrictions are released. Based on these simulations, we conclude that even very long “lock-down” might not suffice if the disease enters a large and tight community (i.e., large household or a care home), where multiple generations of the disease might survive. In addition to that, we reiterate that such strict “lock-down” measures would have significant economic downsides. Furthermore, the implementation requirements of such strict measures in terms of logistics, law enforcement, and provision of the fundamental services would make this scenario impossible.

### E. ICU capacity-based introduction of the NPI can represent an optimal trade-off between the pressure on the healthcare system and the economic damage

One of the interesting strategies to mitigate the impact of COVID-19 outbreak presented in [10] is the adaptive release and introduction of the strict NPI based on the occupation of the ICU capacity. Such “on-off” approach is, in a sense, analogical to the bang-bang control in the classical control theory [54] and utilizes the principles of feedback control theory by selecting the degree of the restrictions based on the value of one of the states, in this case, number of occupied ICU beds. We initialized the simulation as described in Methods section, assuming that, initially, there are no limitations on mobility or mandatory social distancing. We then introduced the simple mobility restriction in terms of the reduction in the number of random contacts to 40% if the total number of 1000 ICU beds was filled to 70%, i.e., the number of cases requiring ICU hospitalization reached 700. These restrictions were released if the demand for ICU beds declined to 50%, i. e., 500 beds (Figure 7). The results show which shows that after the initial long period of the suppression, the ICU-based introduction of the restrictions could keep the demands for the ICU beds below its maximum available capacity. We hypothesize that the ICU-based NPI are particularly effective if the mild restrictions are imposed even if the “Switch ON“ number of ICU is not yet occupied, as these mild NPI can reduce the speed of increase of the ICU cases. The significant advantage of this approach is the potential reduction in the economic and social damage, which is caused by the NPI-related restrictions. However, additional investigation is required to optimize the limits on the introduction and release of the restrictions. The intermittent introduction and release of NPI for the ICU-based restrictions with different bounds is shown in Figure S10.

**Figure 7.**
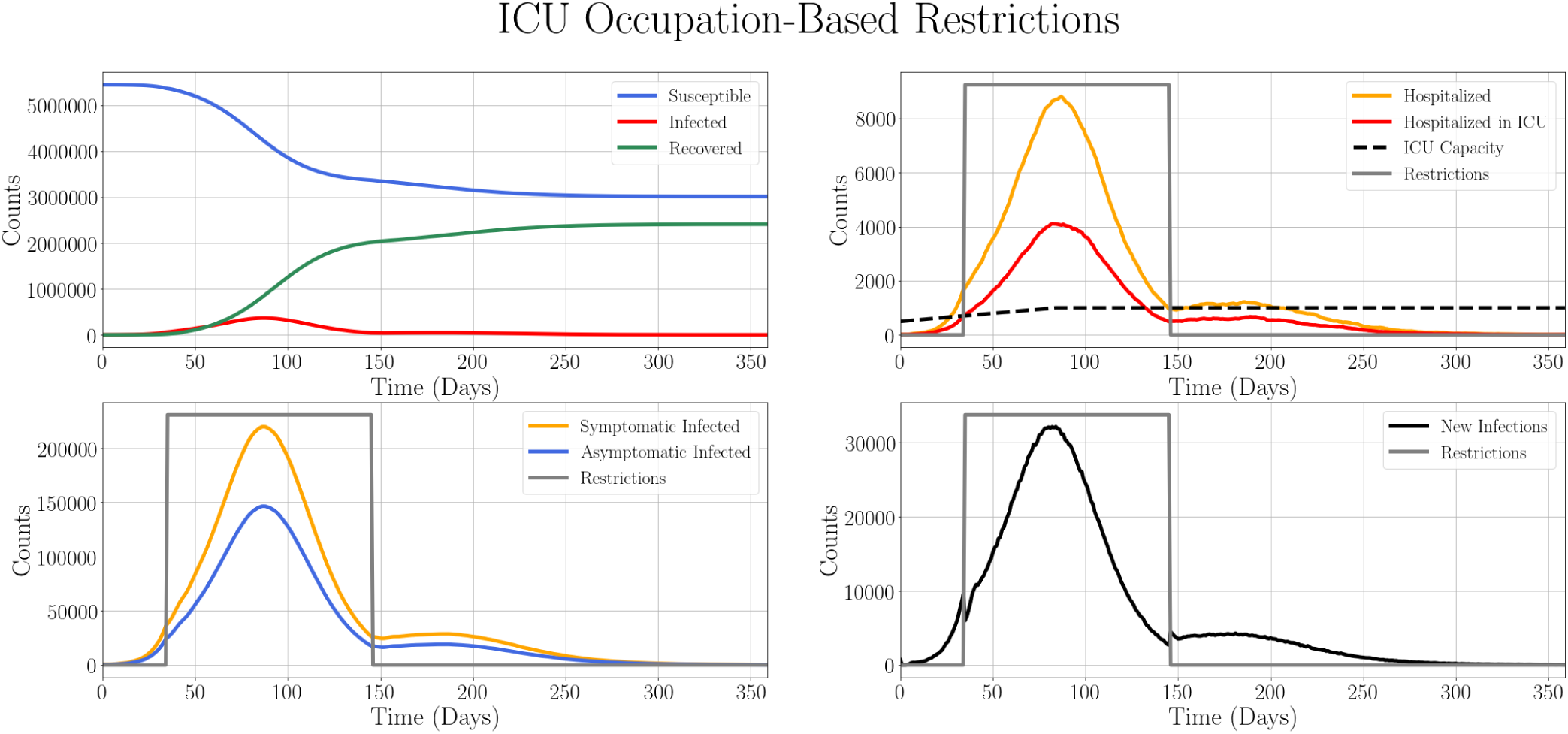
Time evolution of COVID-19 spread through Slovakia in the setting with the strict NPI, based on the relative ICU beds occupation. The restrictions, reduction of the number of random contacts outside the households to 40%, are introduced if more than 70% of ICU beds are occupied and are released when the demand for ICU beds declines to 50% of ICU capacity. (Upper Left) Counts of Susceptible (blue), Infected (red), and Recovered (green) individuals. (Upper Right) Numbers of individuals requiring hospitalization (orange), hospitalization in ICU (red) and the maximum ICU capacity available (dashed black). (Lower Left) The time evolution of the counts of Infected Symptomatic individuals (yellow) and Infected Asymptomatic individuals (blue). (Lower Right) The time evolution of the daily new infections (black). Grey shading of the plots depicts the period during which the strict NPI are in place. Grey line demonstrates the introduction and release of the NPI in terms of mobility reductions.

### F. Superspreading events accelerate the spread of COVID-19

There is a wide discussion of the impact of the superspreading events on the spread of COVID-19 through the population in scientific [15] [55] and non-scientific literature [56]. The superspreading individuals are those causing disproportionately high number of secondary transmissions [55]. Analysis of the impact of superspreading events is very important, as the contact tracing and isolation of the superspreaders can significantly reduce the speed of the disease spread. To evaluate the effect of superspreading events in the population, we assumed that 5% of the infected individuals are superspreaders. For superspreading individuals, we drawn the number of daily random contacts *n_S_* from the Negative Binomial distribution *n_S_* ~ *NB(r* = 15, *p* = 0.1) (Figure S3) [57]. The characteristic number of random daily contacts significantly increased for the superspreading individuals. The number of random contacts for rest of the population, is distributed as *Poisson*(λ = 15) as before. However, to keep the constant effective reproduction number (calculated as in Methods section), the transmission probability in the random contact was decreased to balance the increase in the number of the random contacts. The simulations were run without any NPI. While the steady state values of the states are not influenced by the superspreading, the peak numbers of the infections, hospitalizations and ICU hospitalizations for the scenario including superspreaders are shifted to earlier dates when compared to the absence of superspreading (Figure 8).

**Figure 8.**
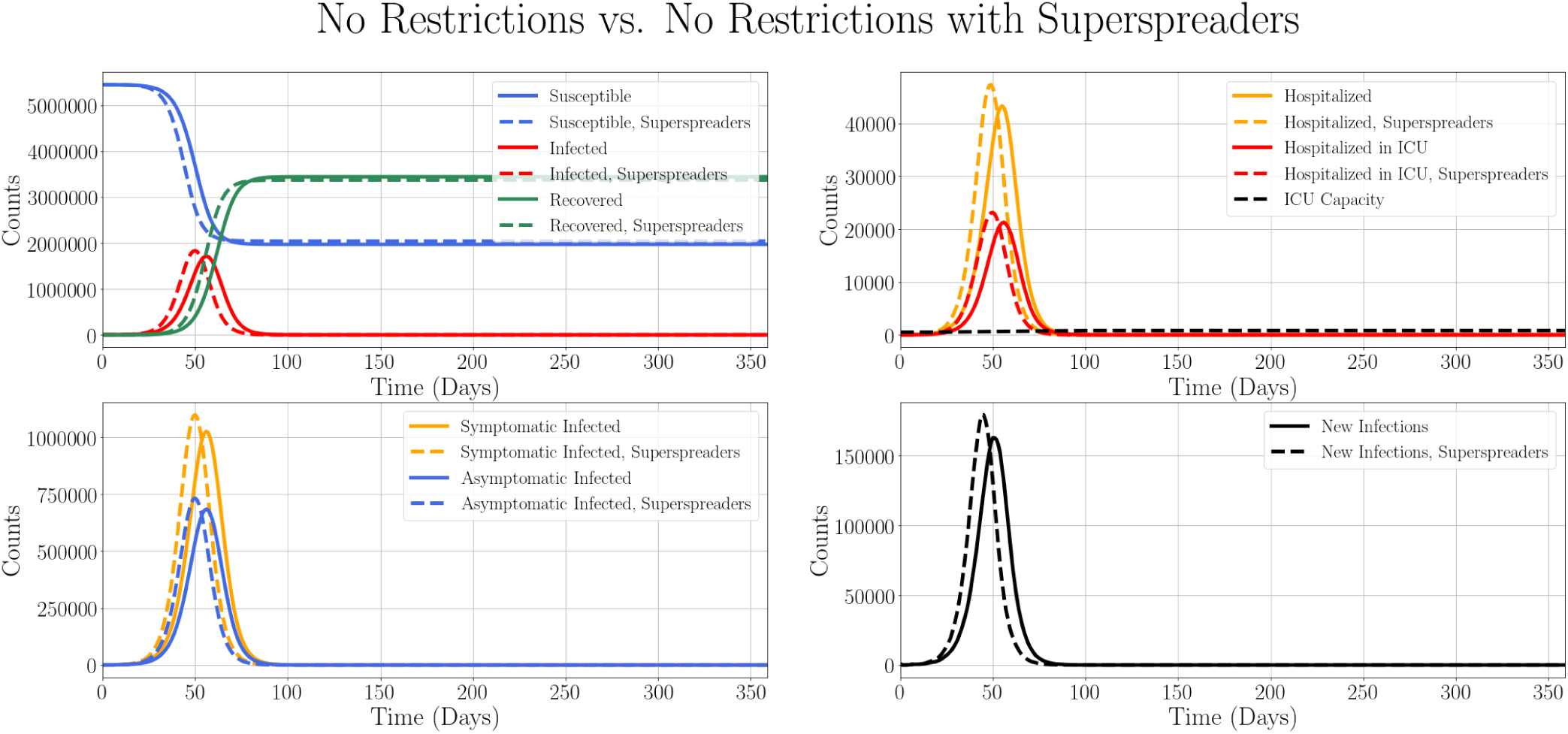
Comparison of time evolution of COVID-19 spread through Slovakia while taking the superspreading individuals into account (dashed line) and without superspreading events (full line). No NPI are imposed in this scenario. (Upper Left) Counts of Susceptible (blue), Infected (red), and Recovered (green) individuals. (Upper Right) Numbers of individuals requiring hospitalization (orange), hospitalization in ICU (red) and the maximum ICU capacity available (dashed black). (Lower Left) The time evolution of the counts of Infected Symptomatic individuals (yellow) and Infected Asymptomatic individuals (blue). (Lower Right) The time evolution of the daily new infections (black).

The key metrics that can guide the selection of the most effective NPI against the disease are the cumulative number of infections, cumulative number of deaths, peak number of hospitalizations, peak number of ICU hospitalizations, and peak number of new infections. We summarized these metrics (Figure 9) relative to the scenario without NPI. The results show the efficacy of the isolation of specific subgroups of the population, in this case, those aged above 60 in mitigation of the negative impacts of the disease, as well as the particular effectiveness of the ICU-based release and introduction of the NPI, even if the further tuning of such feedback control was not performed.

**Figure 9.**
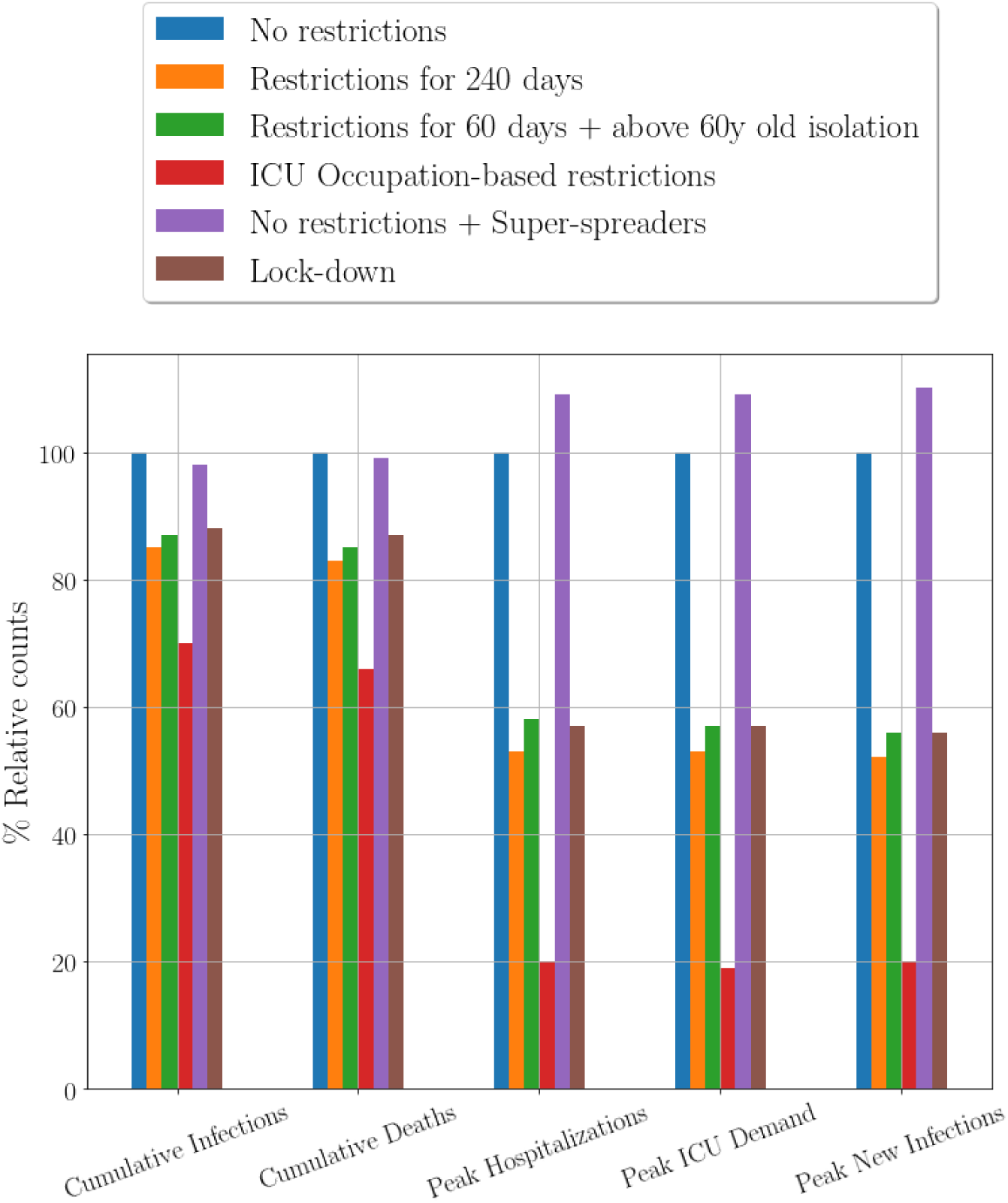
Relative comparison of the cumulative number of infections, cumulative number of deaths, peak number of hospitalizations, peak number of ICU hospitalizations, and peak number of new infections for the different NPI that were utilized in this work.

### G. Analysis of the local spread of COVID-19

The model also allows for the evaluation of the dynamics of COVID-19 for each of the municipalities. This is important for the detailed planning and tuning of the NPI that could limit the spread of COVID-19 by fine-grained local restrictions, utilizing the concept of so-called “smart quarantine“. We show the spread of the virus through Banska Bystrica municipality in the case of no restrictions, strict restrictions, and strict restrictions followed by the social distancing of the elderly (Figure S4).

### H. Evaluation of the decreased household transmission probability and increased initial number of the infected individuals on COVID-19 spread

To evaluate the sensitivity of the model to some of the parameters, we ran the simulations for the decreased transmission probability in the household per day, *β_i_* = 0.032 and for the increased initial number of the initially exposed individuals, which was increased 10-fold in comparison to the value used in the main text, which was *E(t* = 0) = 729 exposed individuals. In the case of the decreased household transmission probability, we maintained the constant reproduction number by decreasing the transmission probability during the random contacts between the individuals (see Methods section). Simulations show that the increase in the initial number of the infected individuals does not increase the total number of the infections; instead it only accelerates the arrival of the peak of the epidemics. The change in the household transmission probability compensated by increased transmission probability outside the household decreases the height of the peak in the number of infections and delays its arrival (Figures S7 and S8). This is in agreement with the assertion that the household transmissions have a strong impact on the disease dynamics.

## Discussion

The spread of COVID-19 is a major public health crisis that requires a decisive, yet measured and targeted response from the public authorities as well as actions from the individual members of the society. This response should be directed by the scientific and clinical facts. Mathematical and computational modelling of the different NPI imposed by the government can significantly improve the adequacy of the policies implemented.

We developed a stochastic, individual-based model of the spread of COVID-19 in Slovakia for the scenario-based projections [20] based on the work in [10] [29] [58]. The model takes into account the clinical characteristics of the disease such as the incubation period, the significant proportion of asymptomatic individuals, and the time required for the progression of the disease from the mild to the severe form that requires hospitalization and subsequent ICU care. We consider the migration of the population within and between municipalities based on the real data [49] and stratification of the population into the age groups and households of different sizes based on the Eurostat data [37]. We utilize the data from Apple Mobility Trends Reports [51] to estimate the reduction in mobility due to the current governmental NPI. In addition to that, this work takes into account the effect of quarantining measures. Inclusion of the distribution of individuals into the households and modelling of quarantining measures represents an improvement with respect to prior modelling work on the spread of COVID-19 in Slovakia [17], which only took into consideration the spatial structure at the level of the municipalities. Moreover, instead of dividing the population into three states (Susceptible, Infected, Recovered), we utilize more sophisticated stratification of the population into up to 8 states. This model allows us to evaluate the impact of NPI on the spread of the disease and effective reproduction number, reduction of which is essential for the eradication of the disease in the absence of the vaccine or herd immunity [13]. The comparison of features of the SIR model with migration presented in [49] and of the model presented here is summarized in Table S1. Based on this comparison, we believe that the model developed in this work and inspired by [10] provides a valuable tool for the *scenario-based projections*.

We stress that the purpose of this model is not to *forecast* [20] the exact number of infected individuals at each time, which is infeasible due to the lack of available data, low level of testing, and necessity to include the more sophisticated structure of the interaction network. Instead, this model allows for the evaluation of the impact of different NPI on the disease evolution when compared to the baseline scenario without any NPI and hence is more suitable for *scenario modelling* [20].

Our analysis shows that the NPI in the form of restrictions on the mobility and quarantining of infected individuals reduce the number of infected individuals and the number of fatalities. This is in agreement with the findings in [49], [10] and [24]. We found that the significant decrease in the contact rates of elderly people (aged more than 60 years) outside their households could have a strong impact on the mitigation of the disease, while allowing for the higher degree of the economic activity. However, the strict social distancing of the elderly might bring some negative consequences in terms of their social well-being [59]. Our analysis suggests that *emphasis on the social distancing of the elderly combined with the short-term social distancing measures utilized by the whole population and ICU-based application of the NPI can significantly reduce the negative impacts of COVID-19* [20].

Furthermore, we analyzed the effectiveness of the so-called “lock-down” scenario, which involves the use of very strict social distancing measures for a limited period of time, which should, in theory, eradicate the virus from the population. Our findings suggest that this scenario is not advisable, as the release of the strict measures would lead to a rapid rise of the infections because the virus could potentially survive in the population that secures the fundamental activities required for the functioning of the society, such as logistics and healthcare. In addition to that, such a scenario would have a devastating effect on the economy and the society.

Our preliminary analysis of the effect of superspreading has shown that inclusion of superspreading individuals sharpens the peak in the demand on the healthcare system and shifts it to the earlier dates when compared to the scenario in which the superspreading individuals are absent. This suggests that superspreading can be an important phenomenon that needs to be captured in the models. Unfortunately, there is currently not enough data that would enable us to accurately evaluate the effect of superspreading. Apart from the lack of knowledge about the number of superspreaders, the probability distributions utilized to simulate the effect of superspreading, such as the Negative Binomial distribution [57], are very sensitive to inputs, such as dispersion parameter [15], and a lack of accuracy in parameter selection can lead to the large variability in the results. Although initial estimates of superspreading parameters were very inaccurate [15], recent research suggests improvement in this accuracy [56] and we hope to utilize it in future work. Additional epidemiological groundwork and contact tracing could improve our ability to adequately assess the effect of superspreading.

We analyzed the effect of the state feedback-like, ICU-based introduction and release of NPI on the dynamics of the disease. In our analysis, this approach has shown promising results, as it was able to reduce the number of cases that require ICU beds below the critical capacity, while potentially allowing for the intermittent release of the restrictions, which would presumably have a salutatory impact on the economy and social cohesion. However, our analysis is by no means complete and further work is required to find the optimal NPI strategy that would strike a balance between the economic and social costs associated with imposing NPI and the potential increased risk to the health of the individuals associated with the relaxation of NPI. Hence, this research direction can be further explored by utilizing the methods of deterministic [60] [54] or stochastic optimal control theory [61], or Reinforcement Learning [62].

Our model has several limitations which can be addressed in future work. First, we need to conduct a detailed sensitivity analysis with respect to the selected probability distributions and their parameters to ensure the robustness of the results against the small changes in their values. Second, more data is needed to inform the selection of several key parameters, such as the number of random contacts per day before and after NPI are imposed. We hope that this lack of data will be addressed by public polls in the near future. Third, while we have done preliminary analysis of the effect of superspreading events, we have not considered the impact of fast local spread in the marginalised ethnic communities living in the eastern part of Slovakia, which can easily become critical sites, accelerating the spread of the disease. Testing and contact tracing in these locations is in progress, and we hope that it will elucidate the potential impact of these communities on the COVID-19 spread. Nevertheless, more data and discussions with experts are needed to fully capture the complexity of this issue. Fourth, currently, we do not have enough data available to describe the interaction among the individuals within the fine social network structure, which prevents us from describing the fine-grained details of the transmission of the disease through the networks of the social interactions among individuals [32] [63]. Instead, we are relying on the random mixing of the individuals within the municipalities. However, we believe that this issue can be ameliorated by the availability of the data from the telecommunication companies. Fifth, we are aware that the effect of *face mask wearing*, which might have a significant role in containing COVID-19 spread, has not been evaluated, as the data on their impact on reduction of reproduction number were published only recently [64]. We are also aware of the absence of consideration of the effect of gloves wearing and relatively closely observed social distancing in the public spaces, which further decrease the rate of disease transmission. Such decrease can potentially be significant and can significantly alter the transmission dynamics. In Table S2 we propose several specific potential improvements that could be implemented in the short and medium-term, along with their implementation period.

## Conclusion

We developed a stochastic, agent-based model that describes the spread of COVID-19 in Slovakia. Our model takes into account the age stratification of the population, distribution of individuals into the household and municipalities, and their migration between the municipalities. Furthermore, the model allows for the evaluation of the local and nation-wide impact of different non-pharmacological interventions, structured social distancing measures and quarantining of the infected individuals on the dynamics of the disease. We supply an easy-to-maintain, simple-to-update open source code that is optimised to run efficiently on a GPU. We utilized this model for the scenario-based projections to assess the effect of various non-pharmacological strategies on the spread of the disease. We believe that it can provide a basis for the informed decision-making of the authorities.

## Data Availability

The code needed to reproduce the simulations is available online on GitHub under GPLv3 licence at the following URL: https://github.com/michal-racko/corona-virus-slovakia. We also developed an independent C++ version of the simulation software. Python and C++ frameworks provided the consistent results. The C++ version of the simulation software can be found at the following URL: https://github.com/dubovsky14/sars cov2 slovakia. Unfortunately, because of the confidential nature of the data utilized, especially the Origin-Destination matrix, we are unable to publish these data online. These data can be obtained upon further request.

## Acknowledgements

The authors would like to thank to Harrison Steel for the useful feedback on the report. Authors also wish to express their gratitude to Maria Bielikova, Michal Kompan, Anton Zajac, Matej Novak, Jakub Kory, Martin Mocko, Peter Zmeko and Robert Moro for the helpful feedback on the work. Authors would also like to thank to Vladimir Boza for his suggestions and discussions. We would also like to thank Martin Smatana, Jan Dudek, and Imrich Berta from the Institute of Health Policies (IZP) of the Slovak Republic for the feedback and for provision of mobility data.

## Code and data availability

The code needed to reproduce the simulations is available online on GitHub under GPLv3 licence at the following URL: https://github.com/michal-racko/corona-virus-slovakia. We also developed an independent C version of the simulation software. Python and C frameworks provided the consistent results. The C version of the simulation software can be found at the following URL: https://github.com/dubovsky14/sars_cov2_slovakia. Unfortunately, because of the confidential nature of the data utilized, especially the Origin-Destination matrix, we are unable to publish these data online. These data can be obtained upon further request.

## Author contributions

M. G. and M. R. conceived the project, developed the model, wrote the code, and evaluated the results. M. R. performed simulations in Python and M. D. performed the simulations in C. M. G. wrote the manuscript.

## Supplementary Information

### S1. Distribution of individuals into households based on the number of inhabitants and age structure

To distribute the individuals into the households in agreement with the data from Eurostat [27] displayed in Figure S1 (top), we formed the probability mass function *p_H_(A, S*), where *A* represents the age interval into which an individual falls and *S* is the size of the household. For every household in each municipality, the number and age of individuals were drawn from the probability distribution with probability mass function *p_H_(A, S*). The simulated results followed the results obtained from the real data as shown in Figure S1 (bottom).

**Figure S1.**
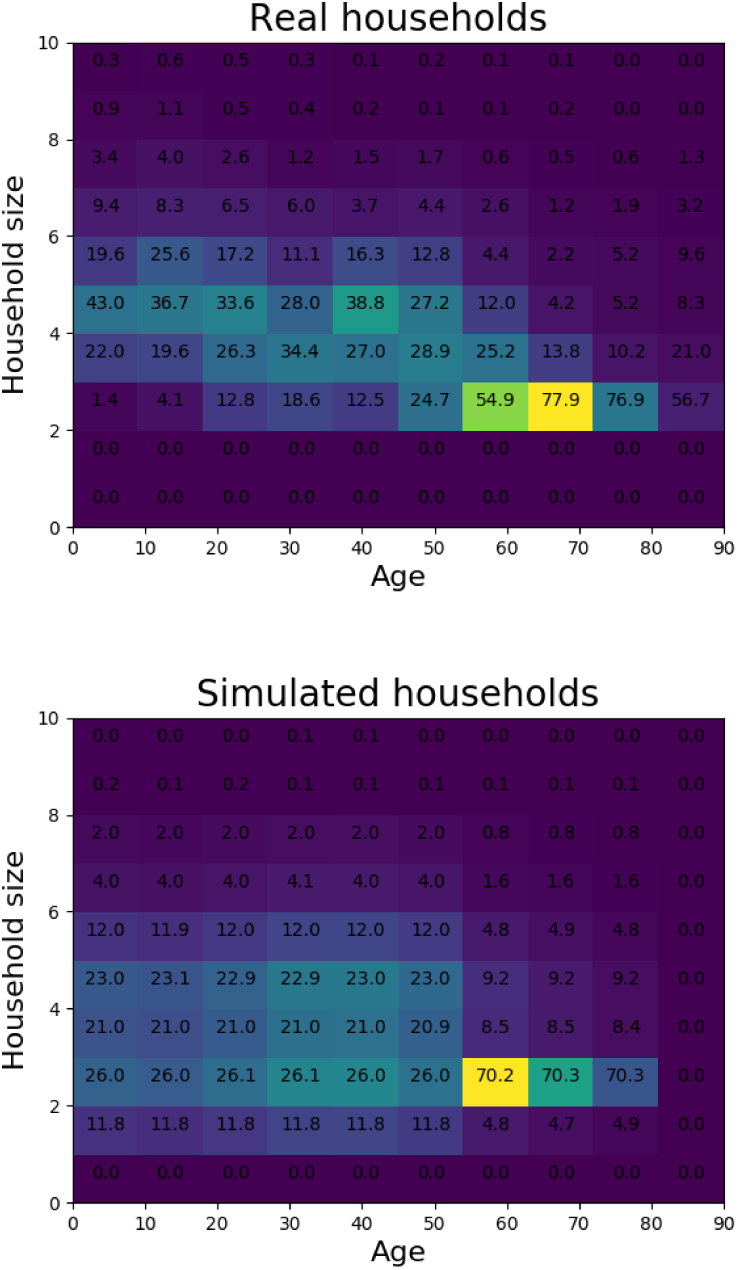
(Top) The typical percentages of individuals of each age living together in each household of corresponding size based on data from Eurostat [27]. (Bottom) The typical percentages of individuals of each age living together in each household based on the simulation.

### S2. Evaluation of the different lengths of the “lock-down” on the disease dynamics

**Figure S2.**
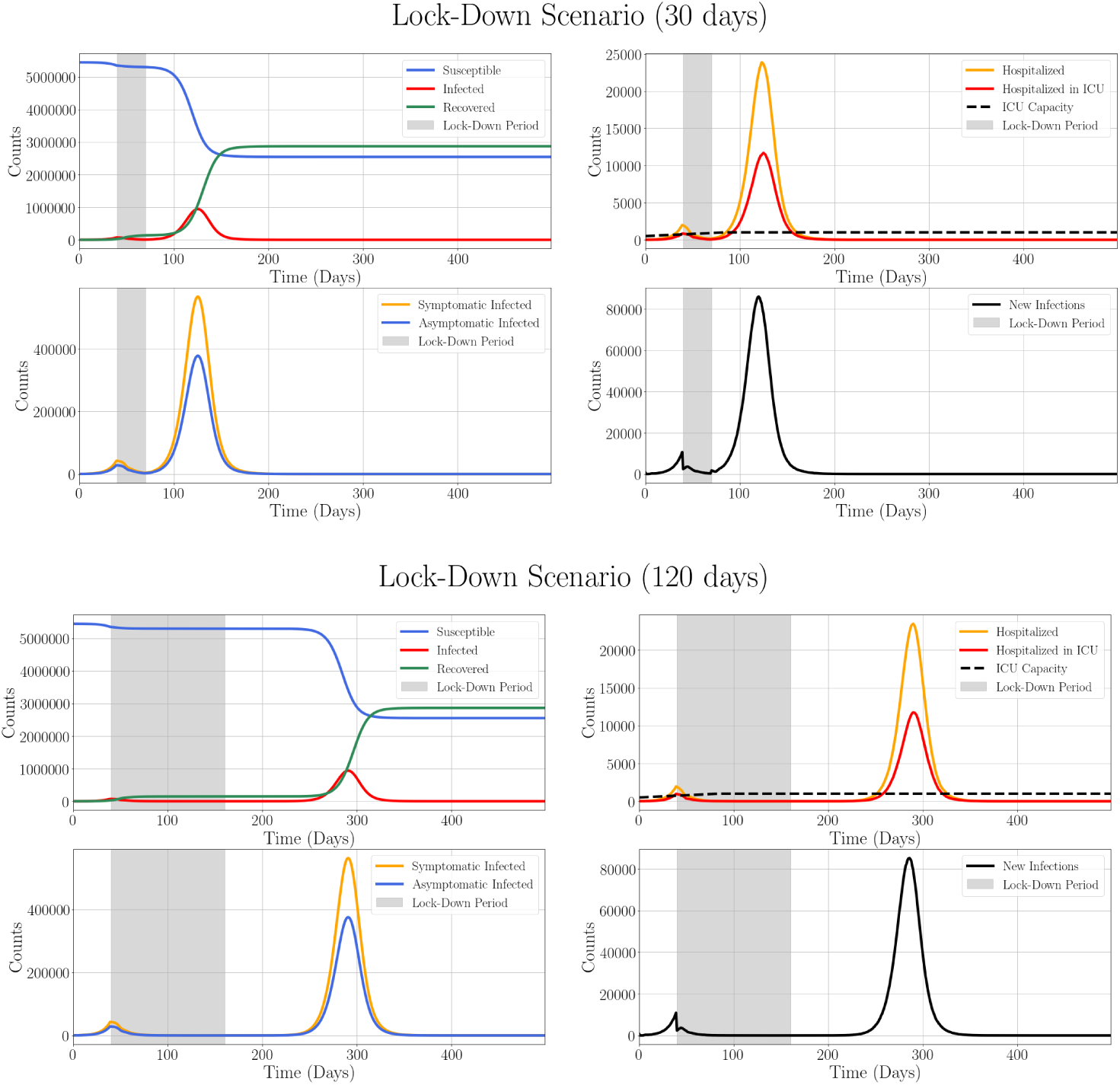
Time evolution of COVID-19 spread through Slovakia in the case of so-called “lock-down” scenario. Initially, the number of random contacts for the individuals travelling between the municipalities is 74% of the non-restricted value. The number of random contacts between individuals is reduced on Day 20 of the simulation (end of April 2020) to 5% of its initial value. (top) Mobility is reduced for 30 days (until end of May 2020) and (bottom) 120 days (until end of August) and subsequently, it is fully restored, while the travel between the municipalities rises back to 74% of its initial value until the end of the simulation. (Upper Left) Counts of Susceptible (blue), Infected (red), and Recovered (green) individuals. (Upper Right) Numbers of individuals requiring hospitalization (orange), hospitalization in ICU (red) and the maximum ICU capacity available (dashed black). (Lower Left) The time evolution of the counts of Infected Symptomatic individuals (yellow) and Infected Asymptomatic individuals (blue). (Lower Right) The time evolution of the daily new infections (black). Grey shading of the plots depicts the period during which the strict NPI are in place.

### S3. Probability distribution of the number of contacts in the case of superspreading

**Figure S3.**
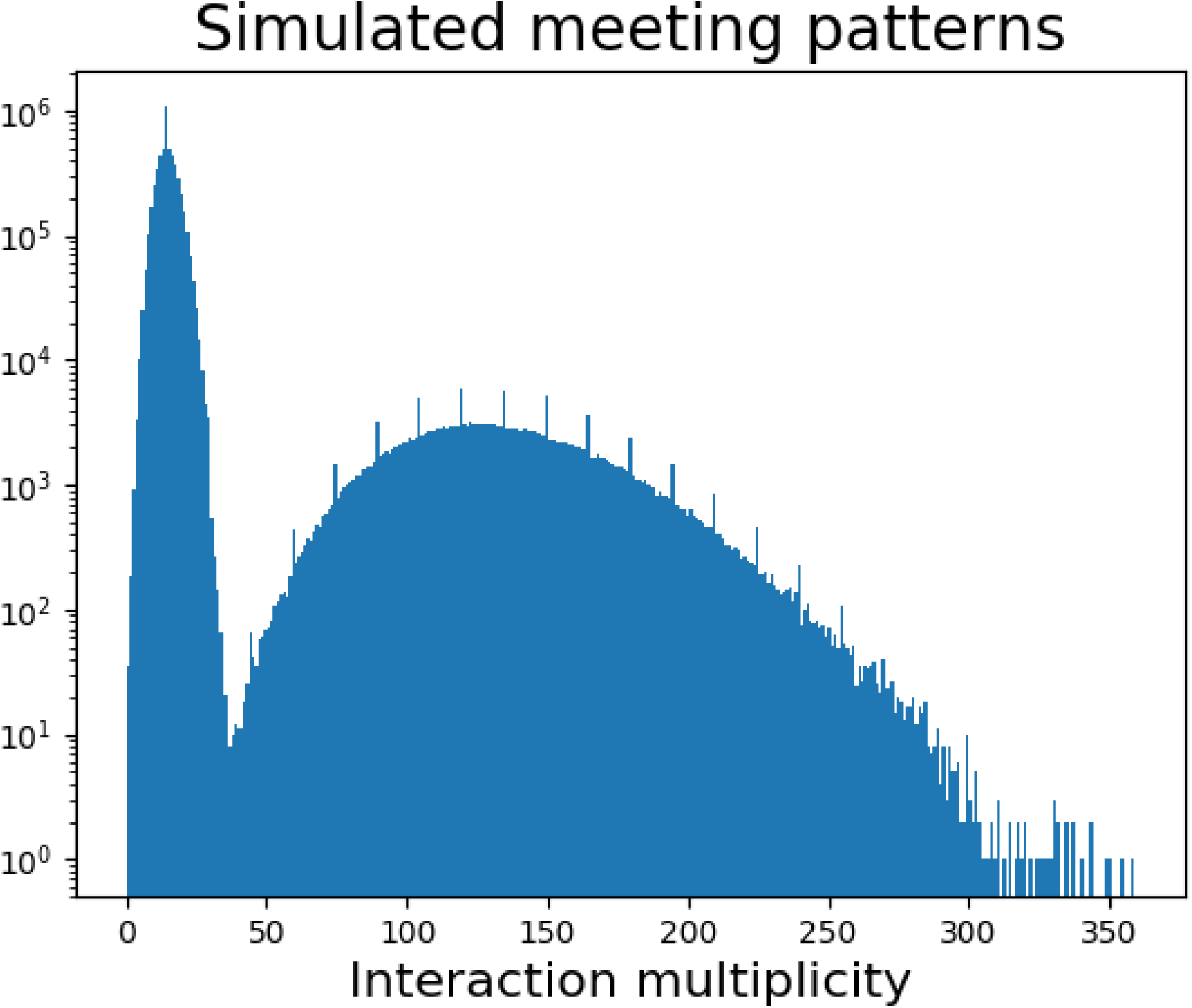
Distribution of the number of random contacts of individuals(x-axis) in the case of inclusion of superspreading and number of individuals with the given contact pattern (y-axis). The numbers of contacts are drawn from the negative binomial distribution *n_S_* ~ *NB(p* = 15, *r* = 0.1) for the superspreaders and from *n* ~ *Poisson*(λ = 15) for the rest of the population. We assume that 5% of the infected individuals are superspreaders.

### S4. Results of the simulations of spread of COVID-19 for a single municipality (Banska Bystrica)

**Figure S4.**
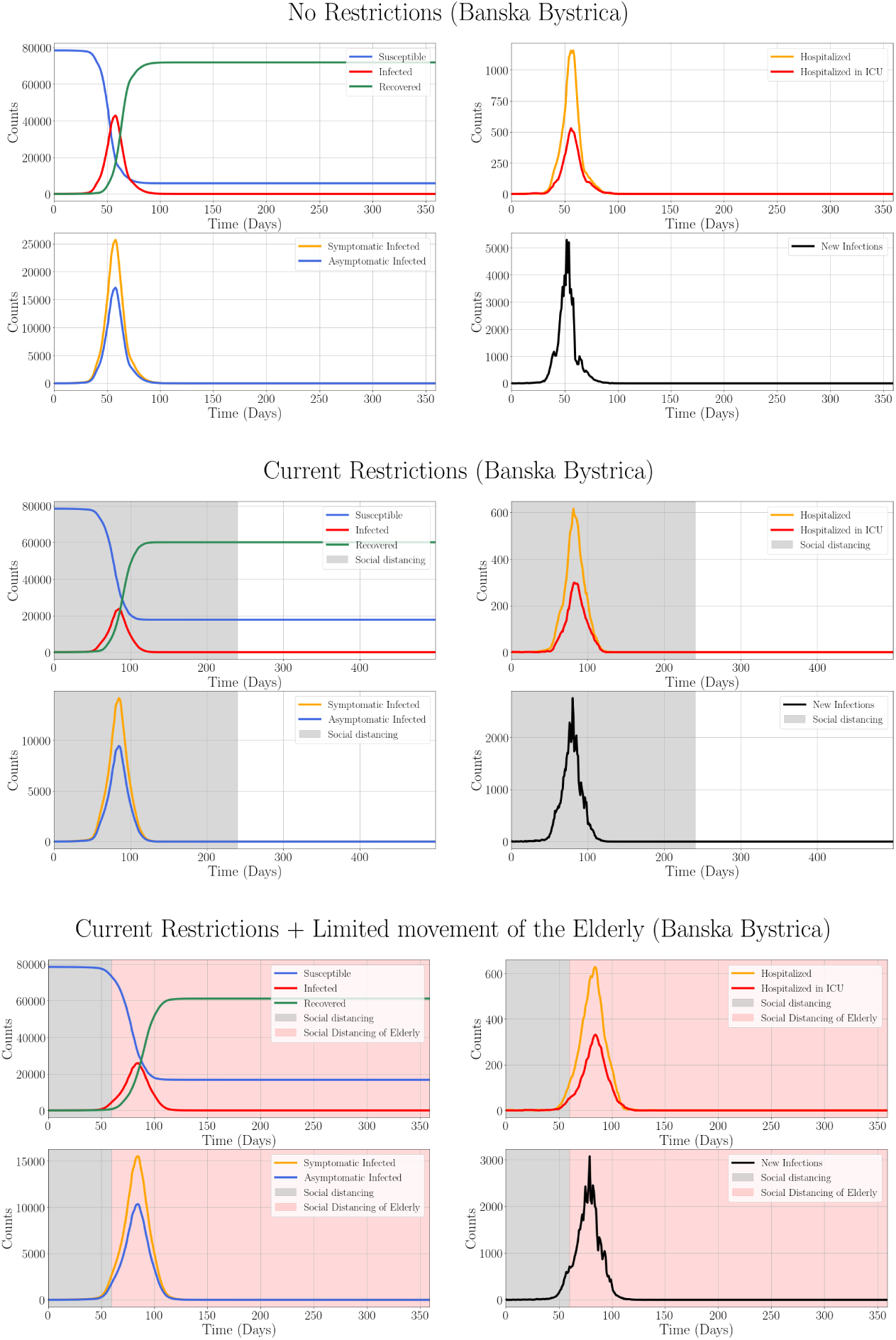
Time evolution of COVID-19 spread through the municipality of Banska Bystrica in the case of no restrictions (top), long-term current restrictions (center), and current restrictions followed by the restriction in the movement of the elderly (bottom). The simulations are conducted as described for the whole Slovakia in the main text. Note, the restrictions are based on the number of ICU occupied in the whole country, rather than in a single region.

### S5. Age stratification of the population and the distribution of the COVID-19-related clinical course time intervals

**Figure S5.**
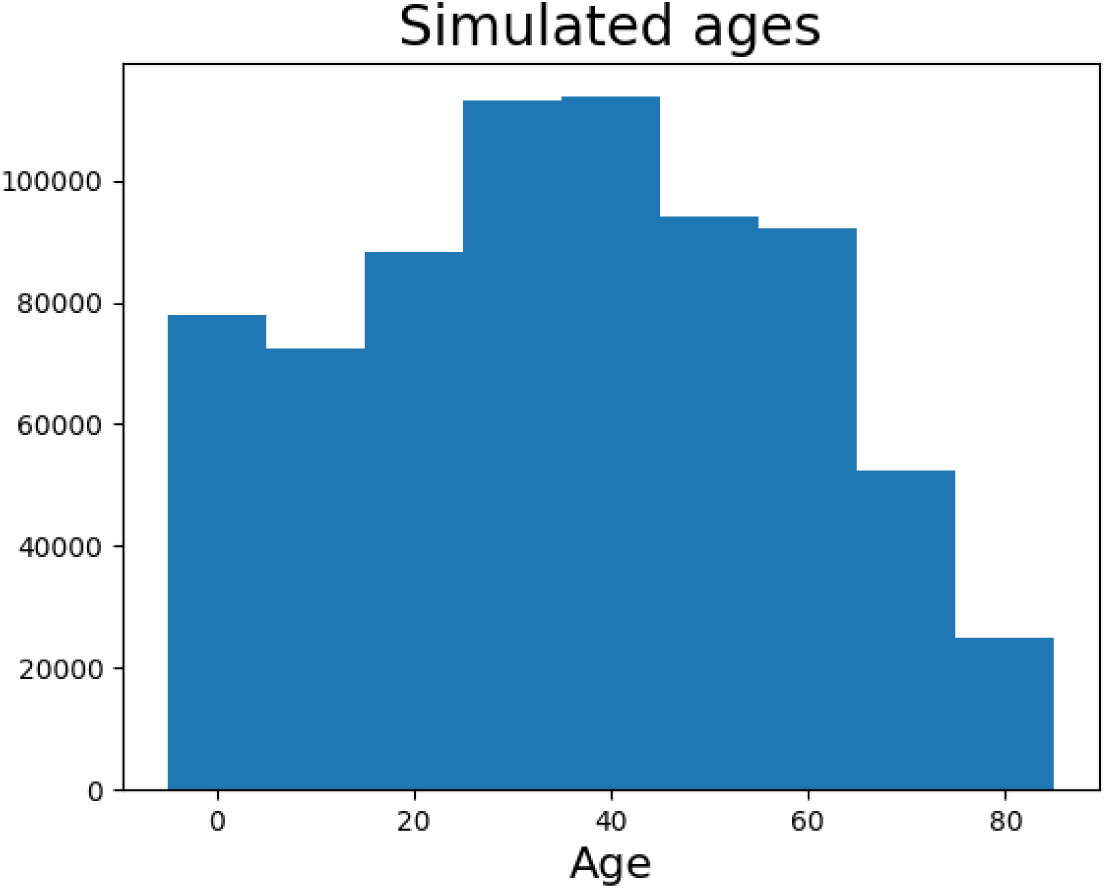
Age stratification of the whole simulated population in the bins of decades (and 80 category).

**Figure S6.**
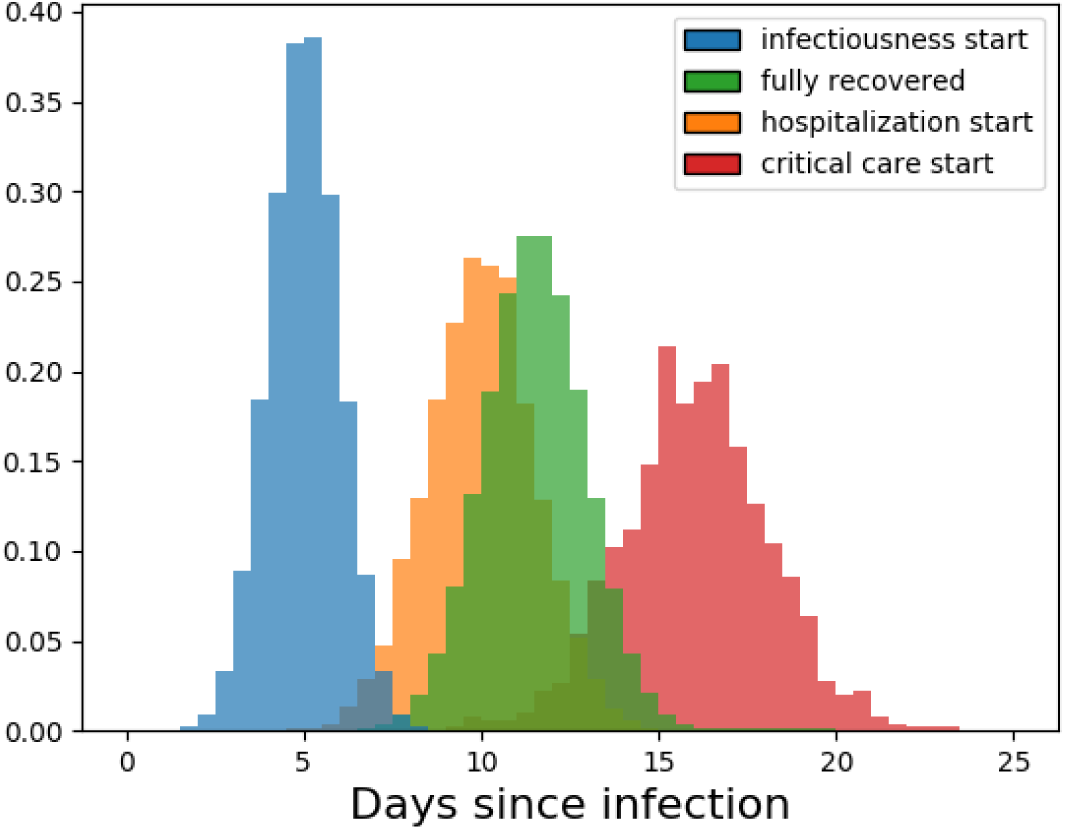
Probability distributions of the periods clinically relevant to COVID-19 disease. After the infection, the patient experiences the incubation period (blue). Subsequently, the patient is infectious (green) and recovered, or hospitalized (yellow) and either fully recovered or moves into the ICU (red).

### S6. Evolution of COVID-19 spread for decreased household transmission probability and increased initial number of the infected individuals

**Figure S7.**
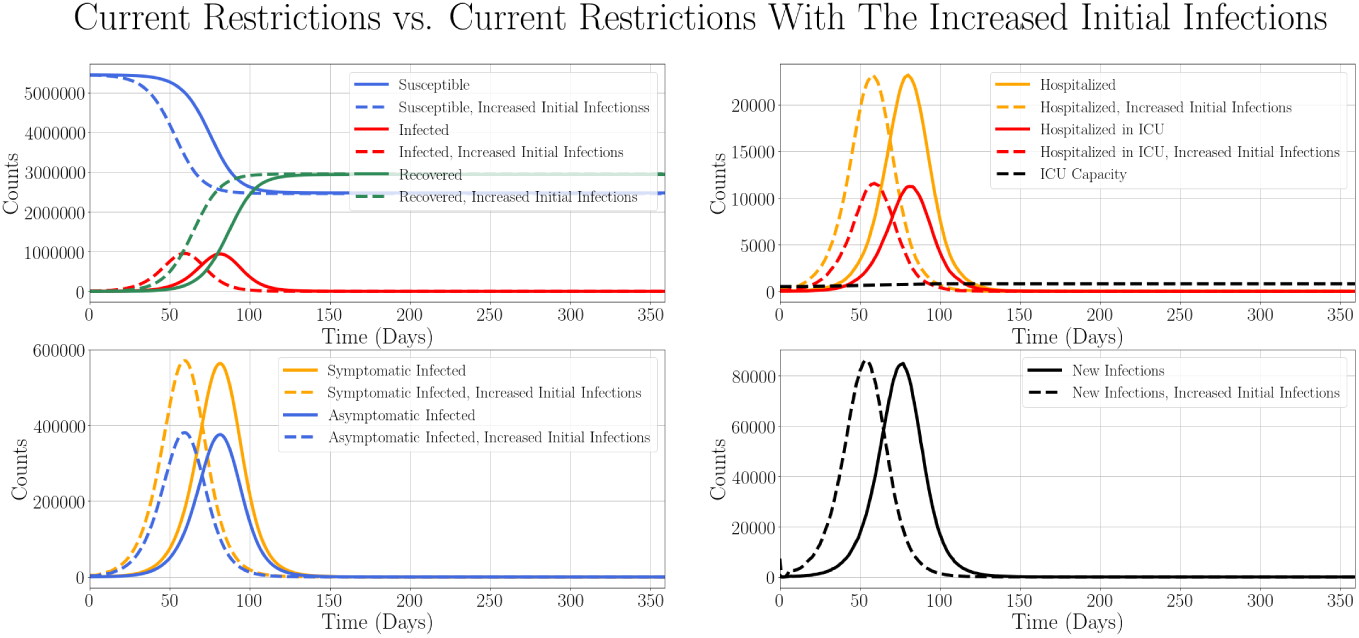
Time evolution of COVID-19 spread through Slovakia for the increased initial number of Exposed 7290 (dashed line), and usual initial number of Exposed 729 (full line). The restrictions imposed are as in Results, subsection B. (Upper Left) Counts of Susceptible (blue), Infected (red), and Recovered (green) individuals. (Upper Right) Numbers of individuals requiring hospitalization (orange) and hospitalization in the ICU (red) and the maximum ICU capacity available (dashed black). (Lower Left) The time evolution of the counts of Infected Symptomatic individuals (yellow) and Infected Asymptomatic individuals (blue). (Lower Right) The time evolution of the daily new infections (black).

**Figure S8.**
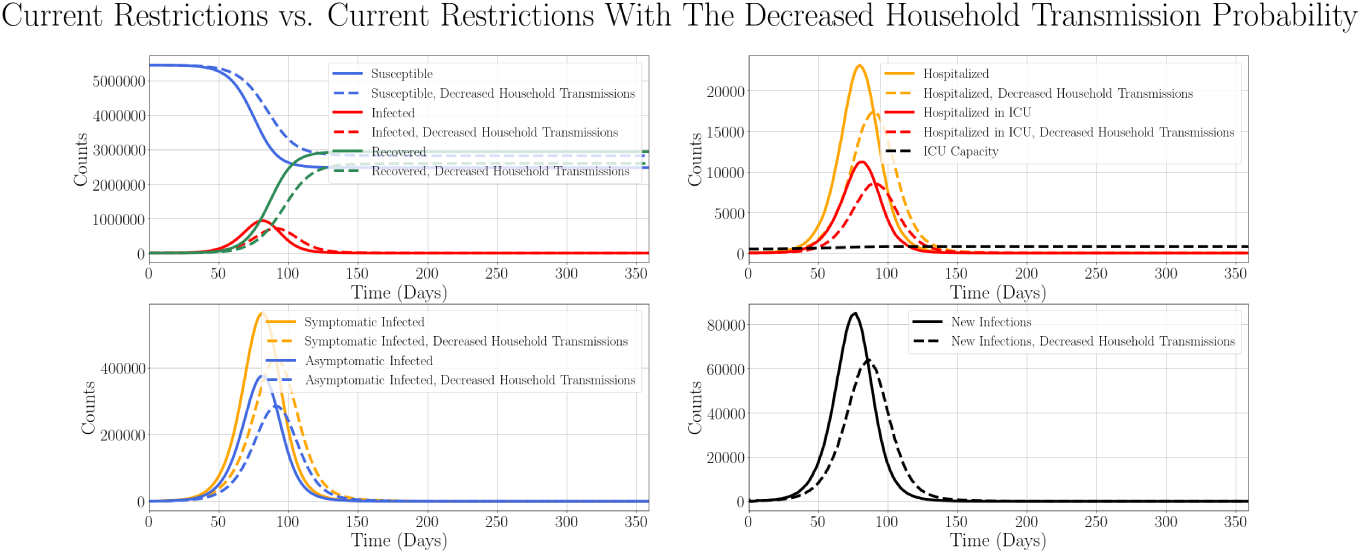
Comparison of time evolution of COVID-19 spread through Slovakia for the household transmission probability reduced to 1/2 of its initial value, *β_H_*_1_ = 0.032 (dashed line) and its typical value *β_H_*_0_ = 0.064 (full line). The reproduction number is assumed to be constant in both cases, i. e. the decrease in the household transmission probability is matched by the corresponding increase in the transmission probability during the random contact. The imposed restrictions are as in Results, subsection B. (Upper Left) Counts of Susceptible (blue), Infected (red), and Recovered (green) individuals. (Upper Right) Numbers of individuals requiring hospitalization (orange) and hospitalization in the ICU (red) and the maximum ICU capacity available (dashed black). (Lower Left) The time evolution of the counts of Infected Symptomatic individuals (yellow) and Infected Asymptomatic individuals (blue). (Lower Right) The time evolution of the daily new infections (black).

### S7. Comparison of the stochastic modelling and deterministic SEIR modelling of COVID-19 dynamics

We compared the COVID-19 dynamics modelled using deterministic SEIR model [13] and stochastic model developed in this work. In the stochastic model, we did not consider the travel between the municipalities or household transmissions, and we matched the reproduction number for both models. The total population was 5,000,000, with 100 exposed individuals on the first day of the simulation. We did not consider any NPI and allowed the disease to spread without any restrictions.

**Figure S9.**
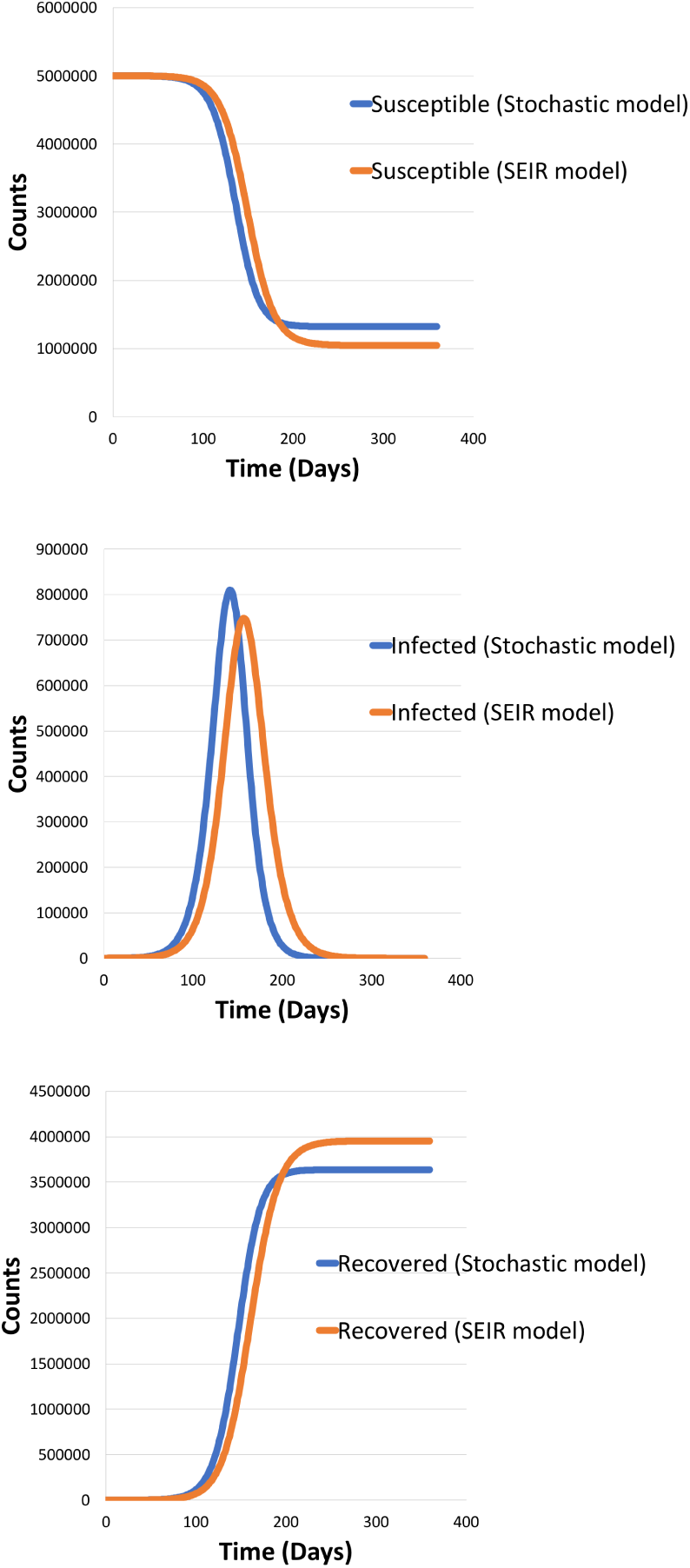
Dynamics of COVID-19 spread in a single municipality using stochastic model (blue) and deterministic SEIR model (orange). (top) Numbers of Susceptible (middle) Infected, and (bottom) Recovered individuals. In the case of stochastic model, recovered individuals do not include the deaths.

### S8. Demonstrating intermittent introduction and release of ICU-based restrictions for COVID-19 outbreak control

**Figure S10.**
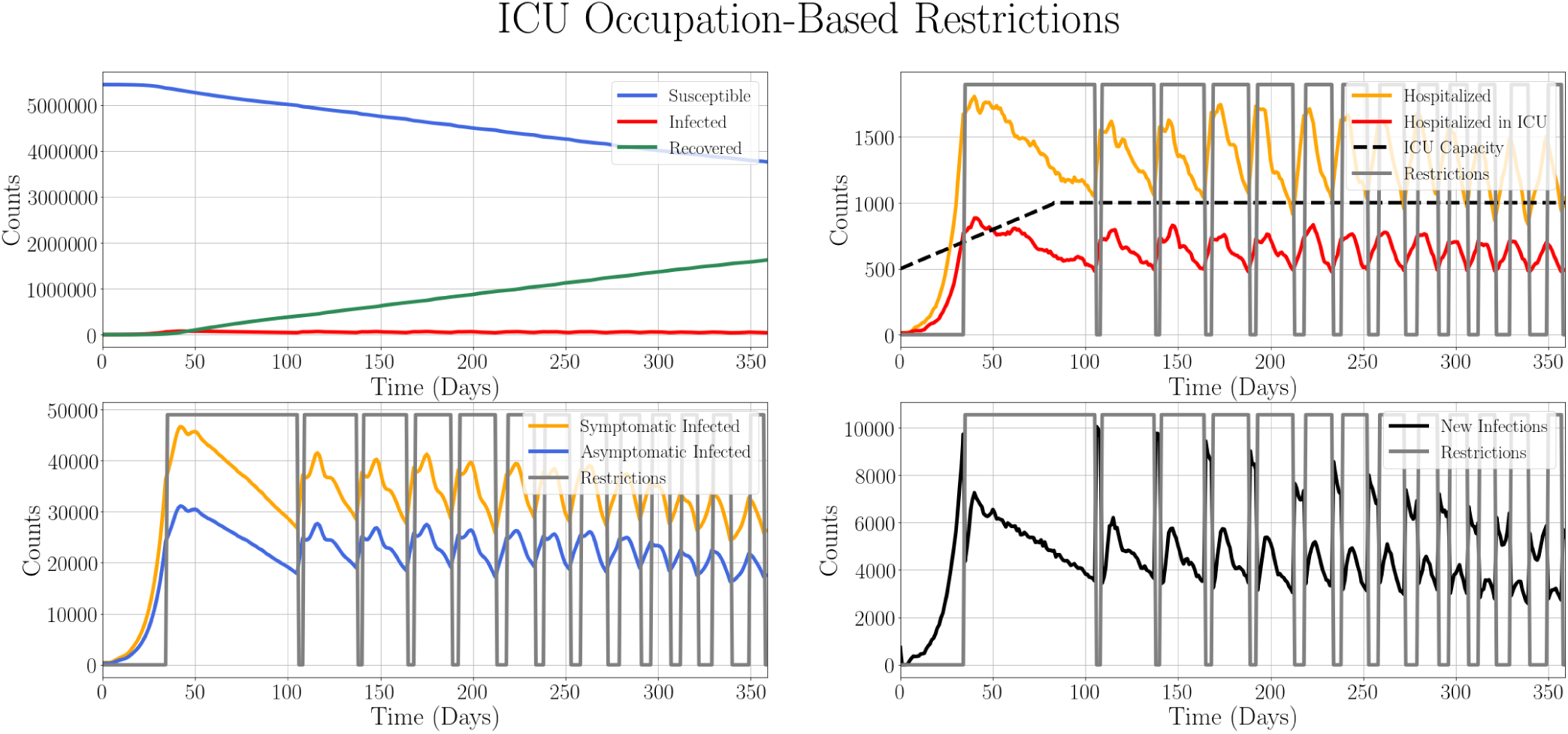
Time evolution of COVID-19 spread through Slovakia in the setting with the strict NPI, based on the relative ICU beds occupation. The restrictions, reduction of the number of random contacts outside the households to 20%, are introduced if more than 70% of ICU beds are occupied and are released when the demand for ICU beds declines to 50% of ICU capacity. (Upper Left) Counts of Susceptible (blue), Infected (red), and Recovered (green) individuals. (Upper Right) Numbers of individuals requiring hospitalization (orange), hospitalization in ICU (red) and the maximum ICU capacity available (dashed black). (Lower Left) The time evolution of the counts of Infected Symptomatic individuals (yellow) and Infected Asymptomatic individuals (blue). (Lower Right) The time evolution of the daily new infections (black). Grey shading of the plots depicts the period during which the strict NPI are in place. Grey line demonstrates the introduction and release of the NPI in terms of mobility reductions.

**Table S1.**
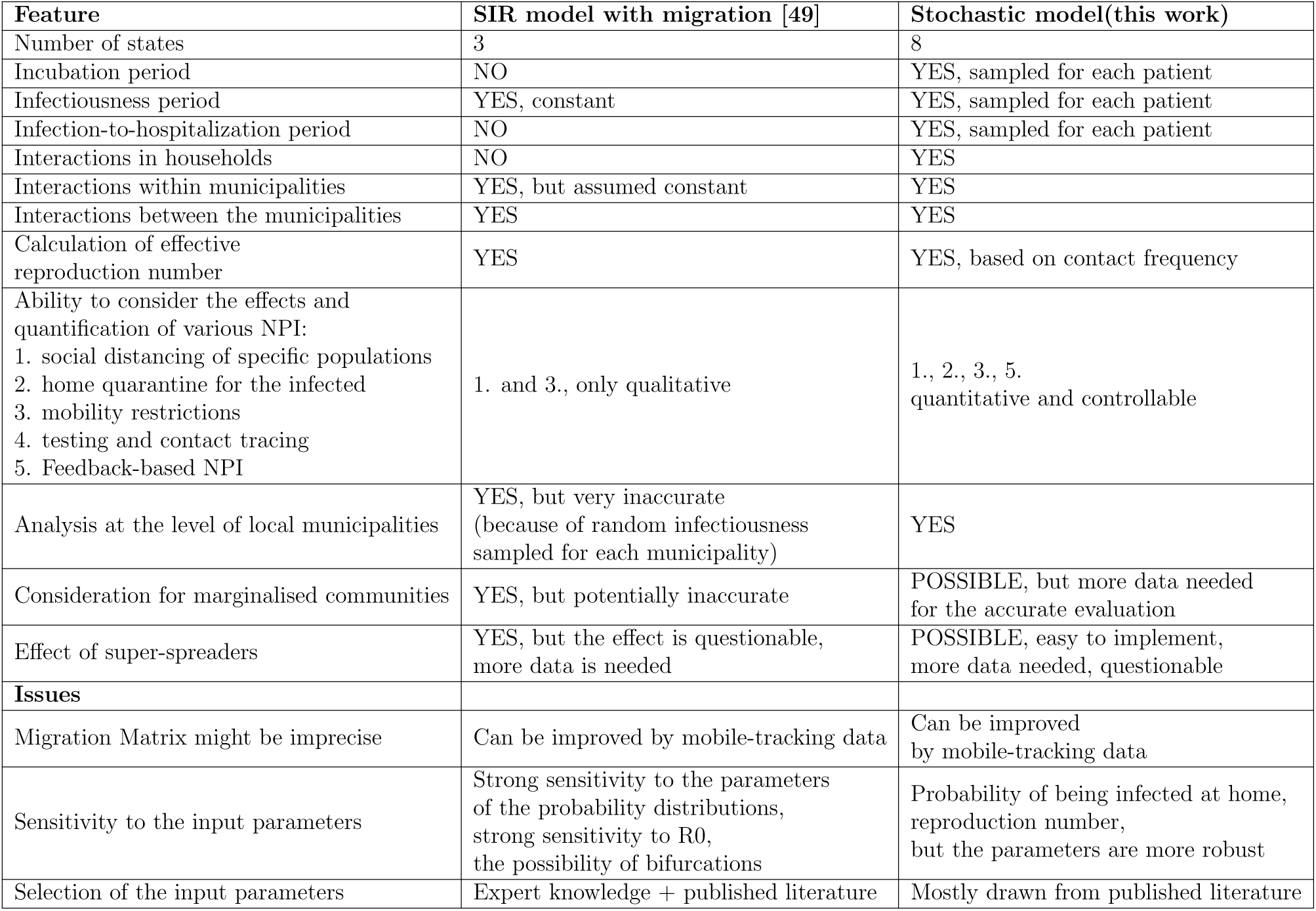
Comparison of the currently used SIR model with migration [17] and the model developed in this work.

**Table S2.** Suggested improvements to the stochastic model that could be implemented in the short-term and the expected implementation period, assuming full-time work of a single person well-acquainted with the code.

| **Suggested stochastic model improvement** | **Expected Implementation Period** |
| --- | --- |
| Robustness analysis of the model and parameter verification | 2-3 man-days |
| Analysis of Marginalised Communities | 2-3 man-days (after the data is available) |
| Evaluation of the impact of COVID-19 spread on the healthcare system capacity in terms of ICU in local hospitals | 1-2 man-days |
| Detailed analysis of the effect of releasing of NPI-related restrictions on Slovak healthcare system | 3-6 man-days |
| Inclusion of the social network analysis in the model to capture human interactions in detail | 5-7 man-days |
| Evaluation of the impact of NPI and restrictions on the economical situation | 7-14 man-days (possibly a team of people) |

